# Human Behavior and Built Environments Shape Influenza Seasonality in the US

**DOI:** 10.64898/2026.08.21.26360922

**Authors:** Giulia Pullano, Andrew Tiu, Jelena Srebric, Donald K. Milton, Linsey C. Marr, Shweta Bansal

## Abstract

Respiratory disease seasonality is widely recognized, yet its mechanistic basis remains poorly understood. Winter environmental conditions have dominated explanations for influenza seasonality in temperate regions, while human behavior’s role has been overlooked. Integrating high-resolution data on influenza cases and human behavior in built environments, we show US influenza seasonality emerges from behavioral adaptations to the environment rather than environmental effects alone. Southeastern US counties maintain persistent summer circulation shaped by weather-driven indoor crowding and built environment vulnerabilities, seeding epidemics as early as August that then spread nationwide. A transmission model incorporating intercounty connectivity, school calendars, and building-modulated transmissibility reproduces this spatiotemporal invasion pattern. These results suggest behavior, both routine mixing and environmental adaptation, shapes influenza seasonality, complementing explanations centered on aerosol persistence or viral survival. Our framework supports epidemic preparedness: modeling southern reservoirs’ connectivity to northern counties, shifting vaccination timing to precede local epidemic onset could reduce influenza burden.

## Introduction

The seasonality of respiratory infectious diseases is widely recognized, manifesting as recurring periods of elevated and reduced transmission. Globally, influenza transmission has been shown to be elevated in cold-dry winters at higher latitudes as well as in hot-humid seasons in the tropics (*1*), a phenomenological pattern whose mechanistic basis remains incompletely understood (*2*). The dominant hypothesis for influenza seasonality attributes winter epidemics to environmental predictors, specifically low humidity and temperature, that enhance aerosol persistence, viral survival and weaken host immune defenses (*3–5*). Yet this account leaves gaps: no comparable mechanistic explanation exists for the hot-humid pattern of transmission, and built environments limit how much time populations actually spend exposed to outdoor environmental conditions.

A complementary but understudied hypothesis attributes seasonality to shifts in host behavior rather than environmental effects on the pathogen alone: human mixing patterns vary with school calendars and holiday travel (*6–8*); human contact and activity fluctuate seasonally (*9, 10*); and human mobility exhibits complex temporal structure (*11*). A growing body of evidence implicates social behavior in influenza transmission broadly: household size shapes viral introduction and spread (*12, 13*); schools amplify local transmission, dampened by holidays (*8, 12*); and mobility networks drive regional spread (*14,15*). Despite this plausibility, models have struggled to integrate behavioral mechanisms empirically, instead imposing phenomenological, time-varying seasonal transmissibility (*16, 17*). Progress in understanding how these social factors, along with related socio-environmental drivers (*18, 19*), shape seasonal patterns has also been hindered by a lack of fine-scale behavioral data spanning geography and time.

Crucially, environment and behavior intersect indoors, where aerosol transmission predominantly occurs and where both aerosol dynamics (*20*) and human mixing patterns vary seasonally (*4, 10, 21*). Yet the physical characteristics of indoor environments and the behaviors that shape exposure within them have received surprisingly little systematic attention in respiratory transmission dynamics (*22, 23*). Ventilation with outdoor air dilutes infectious aerosols and reduces transmission risk (*24*), but rates vary across building types, seasons, and regions (*25*), and extreme temperatures reduce air exchange as buildings seal and recirculate air for heating or cooling (*26–28*). These built environment factors interact with seasonal behavior shifts, as populations spend more time indoors during temperature extremes (*10*), amplifying exposure where ventilation may already be compromised. Together, these factors suggest that geographic variation in seasonal transmission risk arises not only from environmental effects on viral survival (*23*), but from regional differences in how populations interact with their built environments across seasons.

Progress in resolving this gap has been fundamentally constrained by the specificity and spatiotemporal resolution of available surveillance data. Most epidemiological studies rely on syndromic indicators such as influenza-like illness (ILI), which do not distinguish influenza from co-circulating pathogens like RSV, rhinovirus, or SARS-CoV-2 (*29*). Even during peak influenza seasons in the United States, only approximately 30% of ILI is attributable to influenza, obscuring pathogen-specific seasonal patterns, particularly summer circulation masked by other respiratory illnesses. In contrast, laboratory-confirmed diagnoses capture pathogen-specific encounters across diverse healthcare settings, across routine visits, hospitalizations, and screenings, year-round, independent of seasonal expectations. However, public surveillance systems with laboratory-confirmed data remain too sparse (e.g., CDC’s virological surveillance contained, on average, 25 samples per week, per US county during 2024-2025), masking substantial local heterogeneity (*30*). Without spatially granular, pathogen-specific incidence data across healthcare settings, it remains unclear whether influenza seasonality in the US follows a uniform pattern or exhibits regional structure shaped by local behavioral and environmental conditions. A critical need in influenza modeling is thus the ability to empirically quantify and mechanistically integrate spatiotemporal variation in the behavioral and structural factors that drive seasonal patterns of exposure and transmission.

Our study addresses these gaps by integrating high-resolution datasets on laboratory-confirmed influenza incidence, social mixing, and built environment characteristics to characterize influenza seasonality in the US. The US offers a compelling setting to disentangle these drivers, given its broad latitudinal range, diverse climate zones, and social heterogeneity that generate substantial variation in behavior and environmental conditions. We leverage administrative health data to construct pathogen-specific incidence at daily, county-level resolution, including summer months when transmission has remained largely uncharacterized. We pair this with behavioral data capturing spatial variation in transmission opportunities, including air traffic and mobile device data on intercounty mobility, indoor activity patterns, household size and occupancy, and district-level school calendars. To characterize the built environment, we integrate smart thermostat data on climate control as a measure of mechanical ventilation, alongside building characteristics as a proxy for outdoor air infiltration, a form of natural ventilation. Using multivariate statistical analysis and a mechanistic transmission model, we test the hypothesis that human behavior and built environment characteristics shape the spatiotemporal structure of influenza seasonality. Rather than a full account of respiratory disease seasonality, including viral evolution and the complexity of immune history, we isolate how far behavioral and structural exposure factors, alongside vaccination and immunity proxies, explain the spatiotemporal structure we observe.

## Results

### Persistent summer circulation in subtropical regions seeds national influenza epidemics

High-resolution administrative healthcare data measuring confirmed flu cases from 2016 to 2024 reveal that influenza transmission in the United States exhibits strong geographic variation. Contrary to the canonical view of influenza as a winter disease in temperate climates, our analysis shows that influenza transmission persists in the southeastern US during weeks 22–32 (summer in the northern hemisphere). In particular, counties in southern Texas and Florida sustain summer incidence levels that are two orders of magnitude higher than those in northern regions, ranging from 10^−6^ to 10^−3.5^ per capita (Fig. 1a, b). A latitudinal threshold at approximately 30°*N* marks the divide between regions of persistent summer transmission and those where incidence approaches near-zero (Fig. 1a, S4).

**Figure 1:**
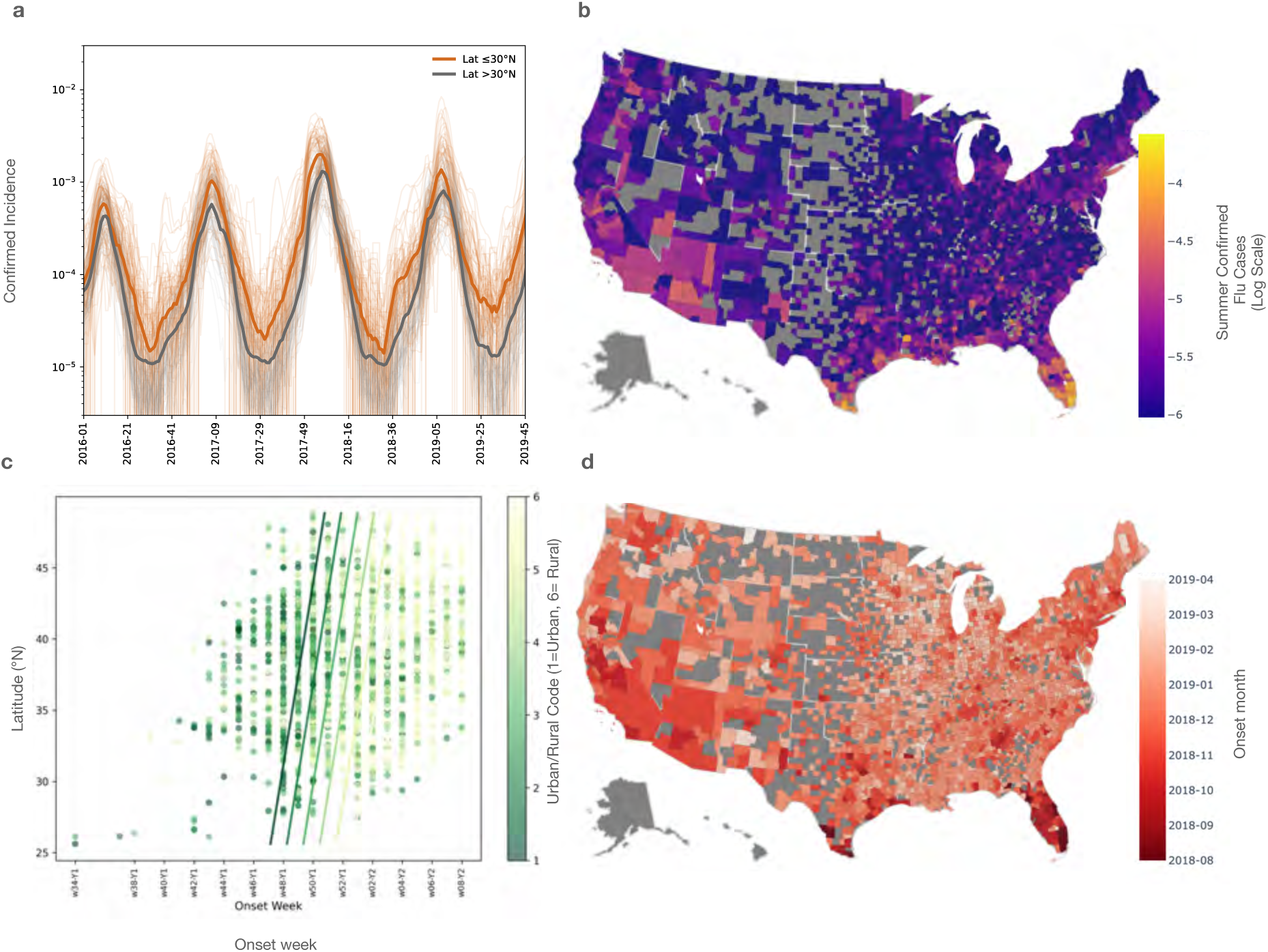
Spatiotemporal patterns of influenza transmission in the United States. (a) Time series of confirmed influenza cases (adjusted and normalized) showing distinct seasonal patterns in locations north (gray) versus south (orange) of latitude 30^◦^N from 2016 to 2019. (b) Map of the spatial distribution of summer influenza incidence (log scale) across US counties. (c) Scatter plot of onset week by county in the 2018-2019 season, plotted against latitude and color-coded by urban/rural classification. Solid lines show OLS regression trends per urban/rural tier, illustrating a systematic earlier onset in urban counties controlling for latitude. (d) Epidemic onset week by county for the 2018–2019 season. Additional seasons are available in the Supplementary Materials.

Beyond sustained summer circulation, we observe early and spatially structured epidemic onset. While CDC surveillance defines the influenza season as beginning in week 40 (early October) our data reveal that epidemic onset in southeastern counties frequently starts as early as August (weeks 34–35) across influenza seasons from 2016 to 2020 (Fig. 1c,d and S2). This early onset progresses northward over eight to twelve weeks, with northern counties typically seeing onset after week 44 (early November). This creates a temporal wave of epidemic emergence moving systematically across the continental U.S. The geographic spread from the south reflects a hierarchical urban-rural structure rather than latitude alone, with major metropolitan areas such as New York and Chicago acting as bridging nodes that receive earlier importation than other northern communities (Fig. 1c, S5). This pattern of propagation aligns more closely with human connectivity than environmental gradients alone, challenging traditional assumptions about influenza seasonality.

These patterns were disrupted during the COVID-19 pandemic (2020–2022) and began to reemerge in the post-pandemic period (2022–2024) (Fig. S3). Notably, similar patterns were observed in independent, out-of-sample wastewater surveillance data collected during the 2023–2024 season (Fig. S10). We also find that respiratory syncytial virus (RSV) exhibits comparable geographic and temporal structure in both summer incidence and onset timing (Fig. S11), suggesting that these dynamics may extend beyond influenza to other respiratory pathogens.

### Behavioral and built environment factors explain geographic variation in influenza seasonality

Using multivariable generalized linear regression models in which all retained, non-collinear predictors were entered simultaneously alongside season fixed effects, we analyzed a suite of county-level predictors (Fig. 2, and described in detail in Materials and Methods). We identify factors associated with summer transmission persistence and epidemic onset timing to reveal that indoor environmental conditions and routine social and regional mixing patterns jointly shape where and when influenza spreads during and after the summer months.

**Figure 2:**
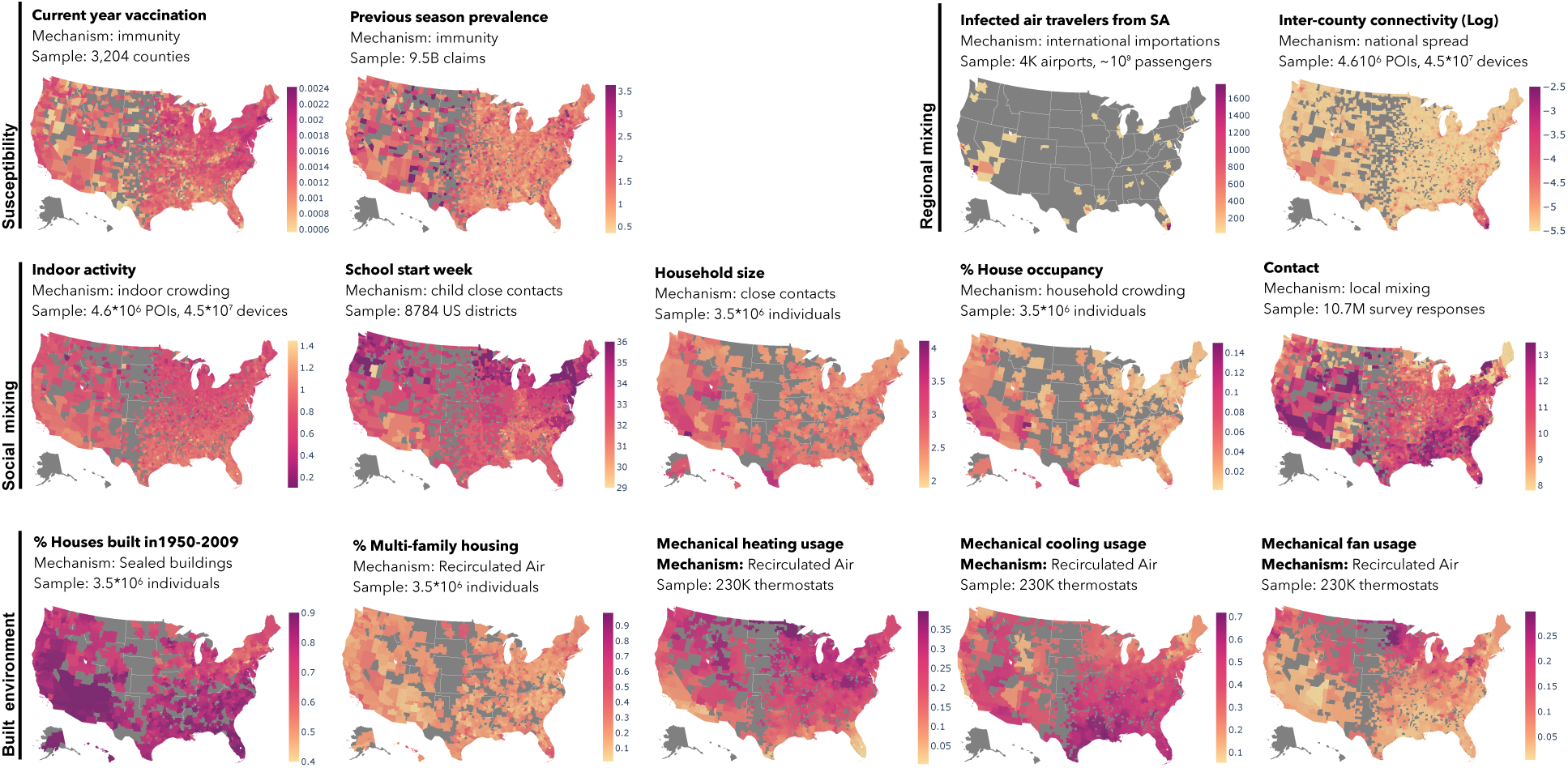
Geographic distribution of hypothesized predictors of influenza transmission. County-level maps showing spatial variation in explanatory variables organized by transmission mechanism: regional mixing (infected air travelers from South America, intercounty connectivity), social mixing (indoor activity, school start week, household size, house occupancy), and built environment characteristics affecting ventilation (mechanical heating, cooling, fan; housing vintage; multi-family housing prevalence). More information in Table S1.

Explanatory variables across US counties show geographic heterogeneity aligned with summer transmission patterns (Fig. 3a; point estimates are reported with 95% confidence intervals). Social Vulnerability Index (SVI), capturing housing vulnerability, was positively associated with transmission (*α* = 0.021 [0.013, 0.029]), consistent with known socioeconomic disparities in influenza burden. The proportion of mechanical fan usage, indicating air recirculation, showed a negative association with summer incidence (*α* = −0.014 [−0.025, −0.003]). Other built environment characteristics were also strongly predictive (housing vintage, a proxy for low infiltration/ventilation: *α* = 0.055 [0.042, 0.067]; multi-family housing prevalence, a proxy for lower ventilation rates relative to single-family houses: *α* = 0.075 [0.067, 0.084]). Routine social and regional mixing were also significantly associated with transmission (contact: *α* = 0.048 [0.042, 0.055]; household size: *α* = 0.035 [0.027, 0.042]; house occupancy: *α* = 0.028 [0.015, 0.040]; population density: *α* = 0.057 [0.052, 0.063]; indoor activity: *α* = 0.023 [0.012, 0.033]). International importation of cases from South America was significantly associated with summer transmission (*α* = 0.025 [0.020, 0.030]). These patterns are consistent with sustained summer transmission linked to crowded indoor settings, elevated ventilation-related risk, and international importations. Results by influenza season are shown in Fig. S9.

**Figure 3:**
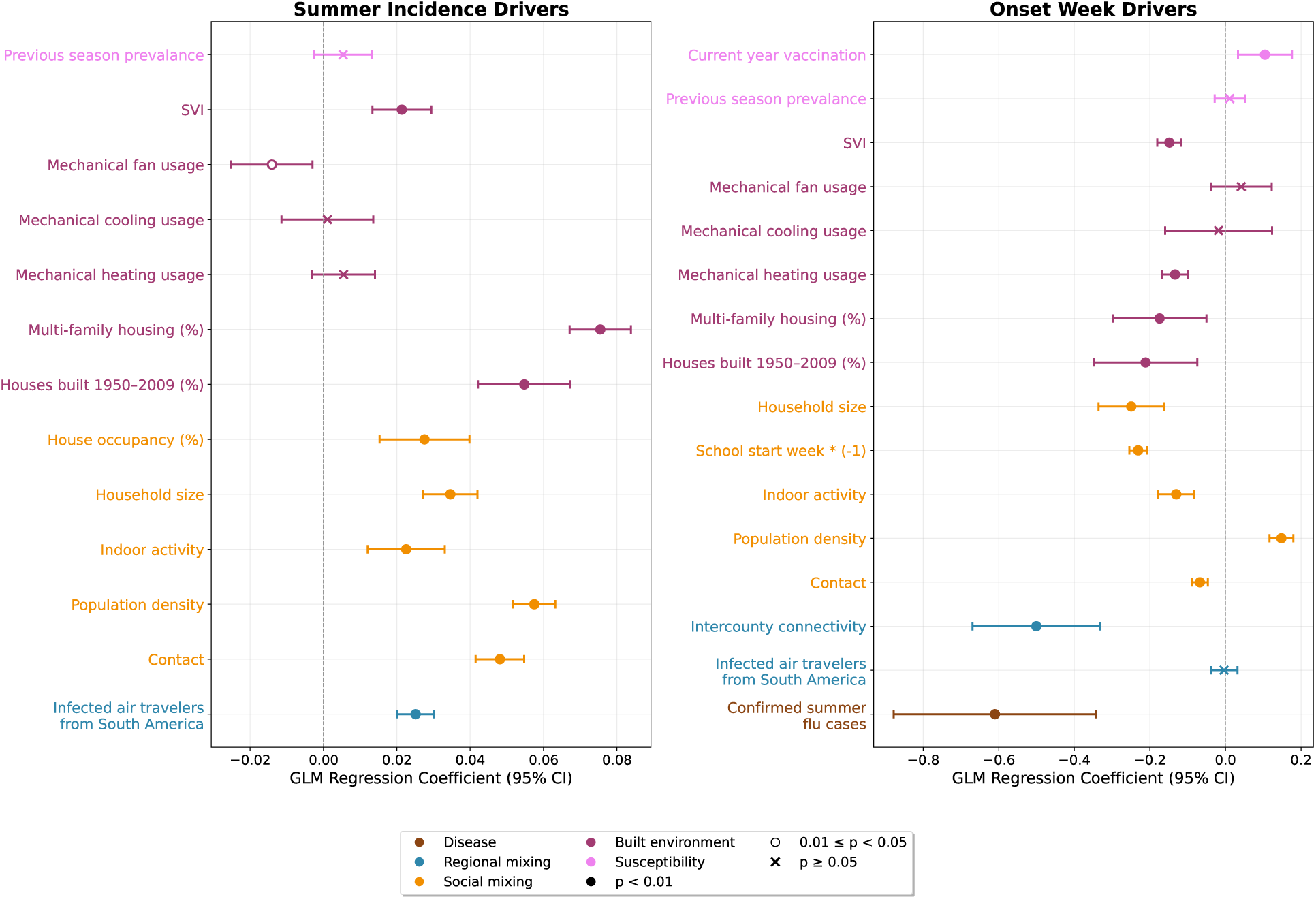
Statistical analysis of factors associated with influenza seasonal dynamics. Standardized regression coefficients and 95% confidence intervals for predictors of (a) summer influenza incidence across U.S. counties, with predictors averaged over weeks 22–32, and (b) epidemic onset timing, with predictors averaged over weeks 29–34 across influenza seasons from 2016 to 2020. Filled dots indicate statistically significant predictors (*p* ≤ 0.01), unfilled dots indicate predictors with mildly significant *p*-values (0.01 ≤ *p* < 0.05), and crosses denote non-significant associations. Confidence intervals were adjusted for spatial and temporal autocorrelation using parametric bootstrap resampling.

Similarly, social and indoor environmental variables were found to explain the geographic heterogeneity in influenza epidemic onset (Fig. 3b). Counties with higher levels of summer transmission experienced earlier epidemic onset (*α* = −0.610 [−0.878, −0.342]), indicating that persistent circulation provides a head start once conditions become favorable for widespread transmission. Highlighting the role of behavior, we found that regional connectivity (intercounty mobility: *α* = −0.501 [−0.670, −0.331]) and social mixing (household size: *α* = −0.249 [−0.336, −0.163]; early school opening: *α* = −0.231 [−0.254, −0.208]; indoor activity: *α* = −0.130 [−0.178, −0.082]; population density: *α* = 0.148 [0.117, 0.180]; contact: *α* = −0.068 [−0.089, −0.047]) jointly determine when and where epidemics emerge following summer circulation. The characteristics of the built environment also determined the timing of epidemic onset (housing vulnerability: *α* = −0.148 [−0.180, −0.116]; mechanical heating usage: *α* = −0.133 [−0.167, −0.100]; multi-family housing prevalence: *α* = −0.174 [−0.299, −0.050]; housing vintage: *α* = −0.211 [−0.348, −0.075]). Results by influenza season are shown in Fig. S9. We include current-season vaccination and previous-season prevalence as a proxy for vaccine and infection-derived host immunity, respectively. (We did not include geographical variability in past vaccination given the waning nature of vaccine-induced immunity.) Current-season vaccination coverage was associated with later epidemic onset (*α* = 0.105 [0.033, 0.176]), whereas previous-season prevalence was not (*α* = 0.011 [−0.028, 0.051]).

We also compared the performance of behavioral and built-environment factors against environmental factors alone. The behavioral model outperformed the environmental-only model for both summer incidence intensity and epidemic onset timing, confirming that behavioral and built-environment factors, rather than ambient meteorological conditions are relevant to influenza seasonality. Full model descriptions and comparison statistics are provided in the Supplementary Material.

Finally, to ensure the robustness of our findings, we evaluated the year-over-year stability of our predictors across the 2016–2019 seasons (Fig. S6). We found that the structural and demographic predictors of summer incidence remain highly consistent across years, whereas predictors of epidemic onset timing exhibit greater interseasonal volatility, reflecting the more stochastic, season-specific nature of outbreak emergence.

Notably, regression analyses on respiratory syncytial virus (RSV) revealed comparable spatial and temporal associations for both summer incidence and onset timing, suggesting that these mechanisms may generalize across multiple respiratory pathogens (Fig. S12).

### Regional connectivity and local transmissibility jointly shape seasonal influenza invasion

To evaluate whether the identified behavioral and environmental factors shape influenza dynamics, we incorporated these predictors into a spatially structured metapopulation transmission model. Covariates with well-defined physical mechanisms linking them to transmission (e.g., mobility, contact, school calendars, and population density) were integrated directly as model parameters. Behavioral and built-environment factors without a clearly specified mechanistic pathway (e.g., mechanical climate control, indoor activity) to transmission were instead incorporated via a second-stage statistical approach, which modulates county-level transmission rates rather than entering through an explicit mechanism (Supplementary Information, Table S5). Starting with observed incidence at week 31 as initial conditions, we then evaluated the contribution of each component to epidemic onset predictions.

The model without spatial coupling (”no connectivity” model) showed poor predictive performance for epidemic timing (Spearman correlation, *ρ*= 0.52, Fig. 4a). In particular, incorporating season-specific vaccination coverage as an immunity proxy improved the no-mobility model from *ρ* = 0.26 to *ρ* = 0.52. Adding intercounty connectivity and contact variability substantially improved model accuracy (*ρ* = 0.63, Fig. 4a), confirming that spatial propagation through human movement networks is essential for capturing observed onset patterns. School calendar effects, incorporated through contact rate changes based on school reopening dates, showed a small but significant improvement in predicting arrival times (*ρ* = 0.64, see additional details in Figure S7).

**Figure 4:**
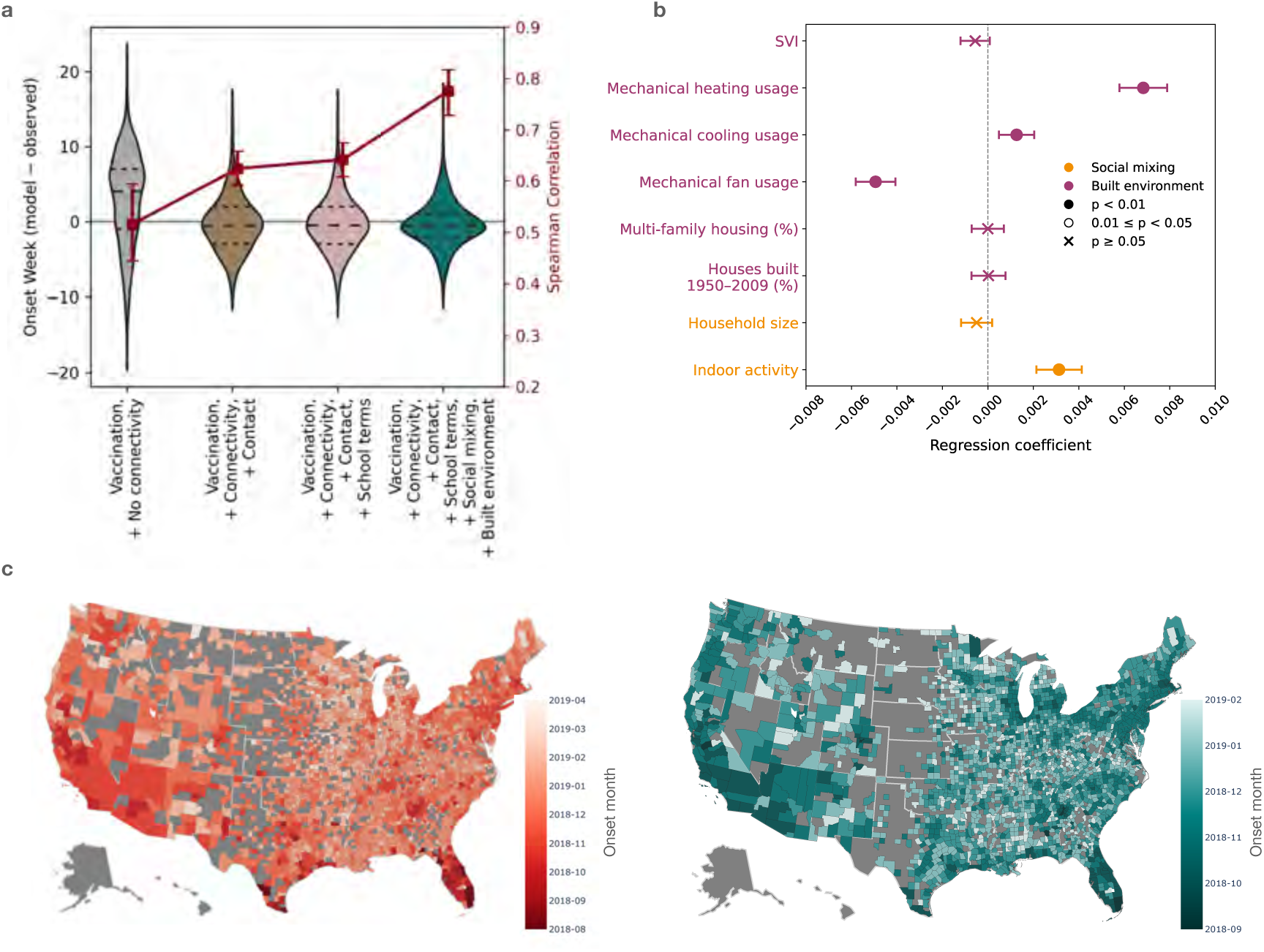
Validation of connectivity-driven epidemic propagation mechanisms. (a) Violin plots show the difference between predicted and observed onset timing for each model configuration with the Spearman correlation coefficient shown in dark red. (b) Linear regression model characterizing factors of residual heterogeneity in transmissibility based on indoor crowding and limited ventilation proxies. (c) Comparison between observed and predicted onset timing. Left) Map of observed epidemic onset timing across the continental United States. Right) Map of model predictions for epidemic onset timing across the continental United States, capturing the characteristic south-to-north progression pattern by the model accounting for connectivity, school terms, and estimated local transmissibility.

To capture remaining variation in local transmissibility, we computed residual transmission rates by comparing model predictions (under the model with connectivity and school terms) with observed data. These residuals, representing unexplained heterogeneity in local transmission intensity, showed positive associations with indoor crowding and proxies of poorly ventilated settings (Fig. 4b) in a Generalized Additive Model (GAM). This model was used to estimate county-level local transmissibility over time. Incorporating local transmissibility modulation based on built environment and behavioral indicators into the spatial model further refined predictions (*ρ* = 0.78).

In Fig. 4c, the heterogeneity in predicted onset times reflects the full model incorporating mobility patterns, school calendar variations, indoor crowding, and proxies of poorly ventilated settings, successfully reproducing the spatial invasion pattern observed in the empirical data.

### Geographically targeted vaccination timing outperforms national rollout

Given the substantial spatial heterogeneity in epidemic timing we identify, a natural policy question is whether vaccination strategies that account for this heterogeneitycould improve outcomes. To address this, we used our parameterized spatial framework, calibrated to the observed timing and coverage of the current US influenza vaccination rollout, as a baseline (Fig. S13). We compared this baseline against two proposed strategies: a uniform national rollout (implemented eight weeks before the observed vaccination rollout) and a geographically targeted onset-based rollout (implemented locally eight weeks before each county’s specific predicted onset time). Since earlier vaccination allows more time for immunity to wane before local epidemics arrive (*31, 32*), we evaluated the impact of waning vaccine immunity on each strategy.

Overall, onset-based targeted strategies outperformed national strategies in total cases prevented relative to the baseline (Fig. 5a). The national rollout is more affected by waning because a uniform shift vaccinates some locations well before their local epidemic onset, giving immunity more time to decline before exposure; the onset-based strategy avoids this by design. Targeting vaccination using model-predicted onset weeks, rather than retrospectively observed onset weeks, yielded comparable results, indicating that this strategy could be feasibly implemented using forecasts rather than requiring observed data in advance. Geographically, the largest gains from onset-based targeting occur in locations with relatively early onset, particularly southern counties and major urban hubs, whose high connectivity otherwise seeds early transmission (Fig. 5b). The two strategies also produce different spatial dynamics: the national rollout preserves the baseline southeast-to-north progression, while the onset-based strategy delays and desynchronizes spread (rank correlation with baseline onset weeks (Fig. S14).

**Figure 5:**
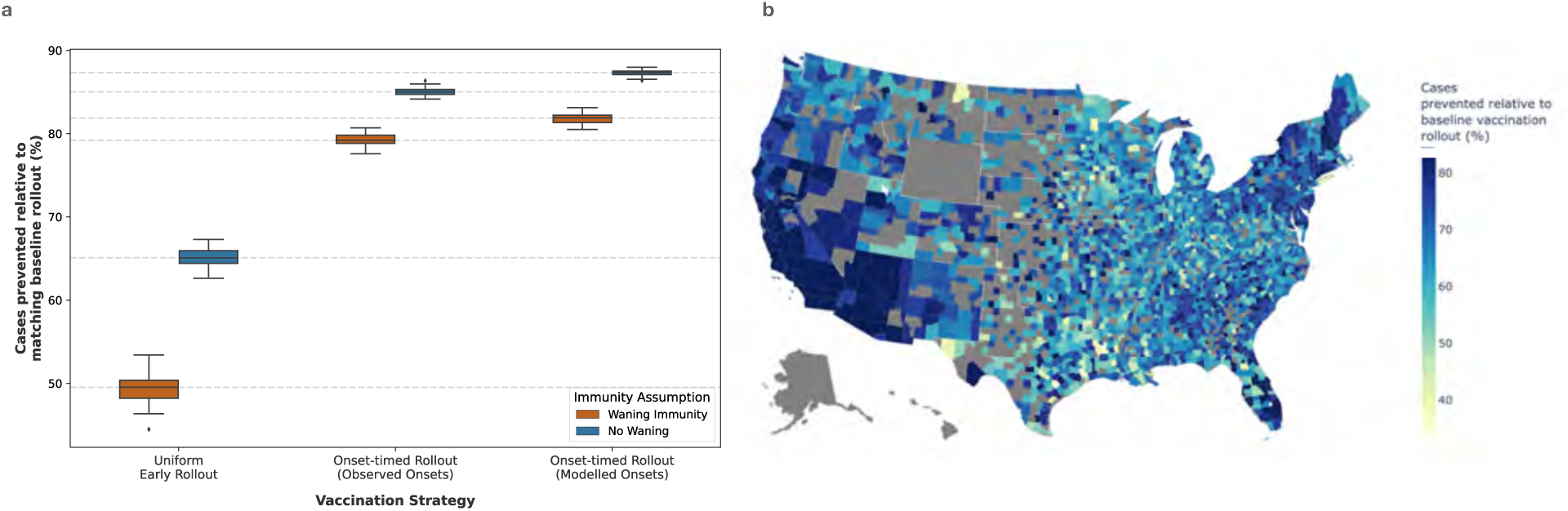
Impact of timing and geographical targeting on vaccination intervention effectiveness. (a) Boxplots displaying the relative percentage of cases prevented across the vaccination strategies: Uniform Early Rollout, Onset-timed Rollout (Observed Onsets), and Onset-timed Rollout (Modelled Onsets). Results are displayed for scenarios modeled both with waning immunity and no waning. (b) Map of the continental United States illustrating the relative percentage of cases prevented at the county level for the Onset-timed Rollout (Modelled Onsets) strategy.

We focus on the 8-week-prior scenarios, since baseline vaccination coverage remains low through roughly the first eight weeks of rollout; 4-week-prior results reported as sensitivity analyses in the Supplementary Material (Fig. S15).

## Discussion

Our analysis reveals that influenza transmission in the United States follows a fundamentally different seasonal pattern than previously recognized. Rather than near-complete extinction during summer months followed by environmentally triggered reemergence, we demonstrate sustained high summer incidence in the southern US, particularly in southern Texas and Florida, with epidemic onsets beginning as early as August and propagating heterogeneously over subsequent months. We observed flu patterns across multiple seasons, including during and after the COVID-19 pandemic disruptions. The post-pandemic reemergence of seasonal patterns aligns with global analyses showing that influenza circulation persisted in tropical regions during the pandemic despite widespread behavioral interventions, highlighting the resilience of seasonal dynamics and providing context for the return of influenza in temperate regions once restrictions were lifted (*33*).

Several factors together help explain persistent summer transmission. International viral importation from South America, supported by recent phylogenetic analyses (*34*), provides seeding into receptive U.S. counties, though importations alone cannot explain the magnitude of summer incidence. Past genomic analyses indicate that viral lineages can also persist season to season (*35*), seeding subsequent epidemics rather than arising from complete reintroduction; this local persistence may further contribute to the maintenance of summer incidence in southern U.S. counties. Once introduced, transmission is sustained by behavioral and environmental conditions: extreme summer heat and humidity in subtropical US regions drive populations indoors into climate-controlled spaces that paradoxically create favorable conditions for transmission, and household crowding compounds this risk by increasing contact and exposure duration. We find that mechanical cooling usage was not a significant predictor of summer incidence intensity, likely because cooling levels are broadly similar across the moderate-to-high incidence counties in our sample; fan usage without heating or cooling, in contrast, is distinctly lower in southeastern counties than in northern ones, making it a more discriminating predictor of summer transmission. Together, these mechanisms reinforce the resilience of seasonal dynamics, with importation, local persistence, mobility, and environmental factors interacting to shape epidemic timing and intensity.

Epidemic onset timing follows a southeast-to-north pattern in the US, which has been observed previously (*15, 19, 36*), but our analysis is the first to demonstrate this pattern using laboratory-confirmed influenza cases across mild and severe disease at fine spatial resolution throughout the US. Persistent summer reservoirs serve as source populations for spatial invasion. Counties with higher summer incidence experience earlier onset, amplifying transmission once conditions favor spread. The timing of this transition is partially modulated by school reopening, which intensifies age-specific mixing among children and accelerates epidemic emergence in counties with preexisting circulation. Weather-driven indoor crowding and poor ventilation rates also shape onset timing. Beyond the early-onset southeastern counties, most locations experience onset in the autumn, when mechanical heating rather than cooling drives indoor climate control. Our mechanistic metapopulation modeling confirms that intercounty mobility networks, rather than latitudinal gradients, govern epidemic spread, with connectivity to early-onset southern counties the strongest predictor of local onset. This finding aligns with recent work demonstrating the key role of human movement in influenza spatial diffusion (*11, 15, 37*). Critically, integrating local transmission heterogeneity, based on built environment characteristics and indoor behavioral patterns, substantially improved onset predictions, demonstrating that these factors are relevant. Future work should aim to establish clearer mechanistic links between these factors and transmission, so that they can be incorporated directly into mechanistic models rather than through statistical proxies.

Several limitations merit consideration. **Healthcare-seeking bias.** Our administrative data rely on healthcare-seeking behavior and may not capture all infections, despite offering unprecedented spatiotemporal resolution across 133 million patients annually; comparison with independent wastewater surveillance data (2022–2024), however, showed alignment with the spatial and temporal patterns we observed. **Ventilation measurement.** Our ventilation measures, mechanical heating/cooling/fan usage and building characteristics, capture two opposing mechanisms, reduced air exchange with outdoor air increasing transmission risk versus dilution and filtration of recirculated air reducing it, but cannot fully capture building-specific airflow; a significant coefficient therefore reflects the net balance of these effects rather than evidence that only one mechanism operates. Direct measurements such as *CO*_2_ concentrations and air exchange rates would help disentangle these mechanisms in future work. **Seasonal variability in built-environment effects.** The predictive contribution of built-environment features varied across seasons, potentially reflecting immunological and antigenic heterogeneity, including strain differences, vaccine mismatch, and atypical seasons such as 2017/2018, not yet captured by our model, an important direction for future work. **Immunity proxies.** We use season-specific vaccination coverage and past prevalence as proxies for population immunity to partially disentangle immunity from mixing and built-environment effects; current-season vaccination proved more informative than prior-season exposure, consistent with immunity waning substantially within a year and being further eroded by antigenic drift (*38*). These proxies cannot capture the full complexity of population immune histories, and more detailed empirical measures are needed in future work. **Seeding and lineage dynamics.** Our mechanistic model, initialized with observed summer incidence, shows how epidemics propagate given southern seeding rather than testing whether this region is the true source of US outbreaks, and establishing clearer causal links between the built-environment factors identified in our regressions and endemic summer transmission remains an open question. Lacking genomic data, we further cannot characterize lineage-level dynamics, such as whether summer introductions from South America persist locally or are repeatedly reintroduced, or whether southern lineages subsequently seed northern outbreaks; our characterization of South American importation relies on phylogenetic evidence from (*34*), and our aggregated case-count analyses are consistent with, but do not themselves establish, this source.

Our fine-scale spatiotemporal findings help resolve the paradox between studies showing low humidity favors influenza transmission in temperate regions (*3, 4*) and field observations of intense transmission in high-humidity US locations (*18*), offering a unified framework bridging temperate and tropical transmission regimes (*1*). This has relevance to other countries spanning broad latitudinal gradients, including Chile (*39*), Brazil (*40*), Australia (*41*), China (*42*), and India (*43*), where similar warm-to-cool propagation patterns have been observed. Our findings also inform understanding of transmission mode: earlier work posited airborne transmission in temperate zones and spray or touch contact transmission in the tropics (*44*), yet aerosol transmission accounts for roughly half of transmission even in subtropical settings (*45*). The generalizability of our findings to other pathogens centers on the relevance of aerosol transmission. Our findings for RSV shows similar geographic patterns in summer incidence, onset timing, and social-mixing associations, though some built-environment predictors diverge in sign or strength, consistent with its partial reliance on aerosol transmission (*46, 47*). In contrast, for pathogens that may be transmitted predominantly via contact (*47*), such as parainfluenza viruses, our built environment factors may plays a far smaller role in their transmission.

Our findings challenge assumptions underlying current seasonal influenza control strategies. Our results suggest that uniform national vaccination timing is poorly matched to the spatiotemporal reality of transmission, and that shifting rollout to align with local epidemic onset, rather than a fixed September-October schedule (*48*), could both prevent more cases and limit the benefit-eroding effects of waning immunity otherwise incurrred. Southeastern communities in particular stand to benefit from earlier vaccination, given their consistently early onset and role in seeding connected regions. We acknowledge that implementing onset-timed vaccination at the county level does face real practical barriers, as coordinating distinct rollout schedules across thousands of counties would strain existing public health infrastructure. We present these results as an illustrative demonstration of the potential gains from onset-aligned timing, rather than a specific policy ready for implementation. Future work should focus on identifying more feasible strategies, such as regional or state-level timing tiers, that still capture the core benefit of aligning vaccination with local epidemic timing without requiring county-by-county precision. Beyond pharmaceutical interventions, our identification of high-risk built environment profiles points to targets for enhanced ventilation standards, portable air filtration requirements, or climate control system upgrades which could reduce transmission risk, potentially across pathogens, increasing the cost-effectiveness of infrastructure interventions.

As climate change intensifies regional temperature extremes and drives populations increasingly indoors, understanding how human adaptations to environmental conditions shape respiratory disease transmission becomes critical. The convergence of rising temperatures, increased urbanization, and climate-driven evolving mobility patterns will fundamentally alter seasonal dynamics (*49*). Integrating built environment characteristics and behavioral responses into transmission models is therefore essential for understanding disease dynamics and developing spatially and temporally targeted interventions that can address respiratory disease burden in an increasingly climate-stressed world.

## Data Availability

All data and code for the project are available at https://github.com/bansallab/flu_seasonality.

## Acknowledgments

We gratefully acknowledge the support and contribution of ecobee and ecobee customers to this research, SafeGraph for sharing mobility data, and Change Healthcare (now part of Optum) for providing claims data and data support. Research reported in this publication was supported by Flu Lab, and NIGMS of the National Institutes of Health under award number R35GM153478. The content is solely the responsibility of the authors and does not necessarily represent the official views of the sponsors.

## Competing interests

The authors declare no competing interests.

## Data and materials availability

Medical insurance claims data used to measure laboratory-confirmed influenza and RSV incidence are proprietary and were provided by Change Healthcare (now part of Optum) under a data use agreement. School calendar data for the 2023– 2024 academic year are publicly available at https://publicholidays.com/. Mobility data from SafeGraph/Advan were obtained under license and are available from Advan Research (https://advanresearch.com/). Air traffic data are publicly available from the Bureau of Transportation Statistics (https://www.transtats.bts.gov/). WHO influenza surveillance data (FluNet) are publicly available at https://www.who.int/tools/flunet. Smart thermostat data were provided by ecobee and are available for research purposes through the ecobee Donate Your Data program (https://www.ecobee.com/donate-your-data/). Weather data from Visual Crossing are available at https://www.visualcrossing.com/. U.S. Census Bureau data are publicly available at https://data.census.gov/. CDC Social Vulnerability Index data are publicly available at https://www.atsdr.cdc.gov/place-health/php/svi/index.html. Wastewater surveillance data are publicly available at https://data.wastewaterscan.org/. All data needed for reproducibility are available at https://github.com/bansallab/flu_seasonality. Code for the transmission and regression models is available at https://github.com/bansallab/flu_seasonality.

## Supplementary Materials

### Materials and Methods

We integrated high-resolution disease incidence data with behavioral and built environment datasets to test whether seasonal behavior and built environment characteristics shape influenza seasonality patterns across U.S. counties. First, we characterized fine-scale spatiotemporal patterns of influenza transmission by extracting summer incidence levels and epidemic onset timing from lab-confirmed influenza case data. Second, we used multivariate regression to identify behavioral and environmental predictors of both outcomes, testing hypotheses including weather-driven indoor crowding, international viral importation, and spatial connectivity as predictors of seasonal dynamics. Third, we developed a spatially-explicit, age-structured metapopulation transmission model to mechanistically integrate these factors and evaluate their causal roles in shaping epidemic timing. Fourth, we used this parameterized model to simulate alternative vaccination strategies based on our onset timing results.

#### Influenza Seasonality Characterization

We analyzed medical insurance claims data (2106–2024) from the commercial healthcare clearinghouse, Change Healthcare, representing approximately 9.5 billion claims from an average of 133 million patients annually. The dataset provides county-week resolution of confirmed influenza cases, covering 40% of county populations across private and public insurers. We normalized disease counts using all-cause denominators: total distinct patients with any healthcare visit at each spatiotemporal scale. A detailed description and validation of this data source are provided in the Supplementary Text.

We extracted two key metrics (Fig. 1b,d): Summer incidence, capturing baseline circulation when transmission is typically low, and epidemic onset timing was estimated from growth rate dynamics.

##### Summer Incidence and Reservoir Classification

Summer incidence was computed as the median weekly influenza incidence rate during the epidemiological low season (weeks 22-32; late May through early August), capturing baseline circulation during the inter-epidemic period. This window was defined to capture the off-season influenza baseline following the end of the preceding flu season (week 20). Week 22 corresponds to approximately June 1, the conventional start of meteorological summer. The window closes at week 32, two weeks before the earliest observed epidemic onset across all study years (week 34), ensuring the summer incidence metric reflects pre-epidemic conditions and does not capture any early epidemic signal.

Using this metric, we defined summer reservoir counties as those sustaining continuous influenza circulation through the low season, operationalized as maintaining a median weekly incidence above the national baseline during weeks 22-32.

##### Epidemic Onset Timing and Bridging Hub Classification

Epidemic onset timing was estimated from weekly growth rate dynamics. We identified candidate onsets when the instantaneous growth rate exceeded *r* > 0.1 for at least three consecutive weeks. Case counts below 8 were set to zero before growth rate calculations to reduce volatile fluctuations associated with low sample sizes. We defined hub counties as highly connected urban counties that receive early viral importation—exhibiting epidemic onset in the earliest quartile for their respective latitude band—but do not sustain baseline summer incidence above the national threshold (i.e., non-reservoirs). To evaluate the relative contributions of geography and urbanicity to onset timing, we evaluated county-level onset across all four seasons as a function of latitude and the NCHS Urban-Rural (U/R) Classification scheme (codes 1-6). We fit Ordinary Least Squares (OLS) regression models predicting onset week from standardized latitude and U/R status (*n* = 8, 752 county-season observations). Continuous predictors were *z*-standardized to compare standardized effect sizes. We additionally evaluated U/R status both as a continuous gradient and with season fixed effects, testing for potential latitude - urbanicity interaction effects. Full regression analysis is provided in the Supplementary Material.

#### Data Sources and Hypothesized Seasonal Predictors

To systematically test hypothesized mechanisms underlying influenza seasonality, we assembled a comprehensive dataset linking built environment characteristics and social and regional behavioral patterns. We organized potential factors into three categories based on their hypothesized mechanisms (Fig. 2): regional mixing, social mixing, and built environment conditions that might affect ventilation Comprehensive information on all datasets used in this study is provided in Table S1 and details on all datasets are provided in the Supplementary text.

##### Regional Mixing

We hypothesized that both international travel and inter-county mobility within the United States shape the spatial and temporal structure of influenza seasonality through a two-stage process: initial international seeding followed by domestic diffusion. We quantified importation pressure from international destinations by integrating WHO surveillance data (2106–2019) (*50*) with air passenger traffic data (*51*), estimating time-varying infection rates among arriving passengers and allocating them to destination counties based on airport proximity and population weights. Using this approach, we found that the majority of importations in US from May to October arrive from South America (Fig. S1). This is consistent with recent phylogeographic work (*34*) which has shown that direct seeding of US epidemics during April-September originates in South America (in contrast to past work which proposed Southeast Asia as the primary evolutionary source of H3N2 influenza variants (*52, 53*)). We then used this metric to inform the multivariate regression and characterize the contribution of importations to sustain summer incidence and seed onset.

For domestic diffusion, previous studies have demonstrated that disease spread across the US depends critically on mobility networks (*14, 54*). We captured domestic connectivity using SafeGraph data (2019–2021, over 45 million devices) aggregated to county-level daily visit networks (*54*). This enabled us to quantify how inter-county mobility propagates infection from early-onset, high-summer-incidence counties to connected regions, synchronizing epidemic timing across the spatial network.

##### Social Mixing

For respiratory pathogens such as influenza, transmission risk is amplified in enclosed environments where aerosols accumulate and ventilation might be limited (*45*). Household and non-household interactions might intensify close-contact frequency, increasing both the probability of viral introduction and the persistence of within-household transmission chains. Increased indoor activity brings susceptible and infectious individuals into closer, prolonged contact in spaces with reduced air exchange, facilitating sustained viral exposure even at moderate contact rates. Climatic extremes (particularly cold winters and hot summers) further drive people indoors, strengthening this behavioral-environmental coupling. Social mixing was characterized through four complementary mechanisms:

- Non-household contact was captured using county-level, non-pandemic baseline contact rate estimates derived from the US COVID-19 Trends and Impact Survey (*55*), to capture geographic variability in overall social mixing.
- Household crowding was assessed using U.S. Census Bureau (American Community Survey) data on occupancy and household size (*51*). Occupancy was captured by the number of persons per room on average.
- Indoor activity, school calendars, and household crowding. We quantified county-week variation in indoor activity using an indoor activity index measuring the propensity for visits to indoor versus outdoor locations, derived from around 4.6*10^6^ times points of interest classified by primary setting type (*10*).
- School calendars from public school districts (2023–2024 academic year) served as a proxy for shifts in child contact patterns (*56*), capturing the synchronized return of students aged 5–17 to dense classroom networks. We assumed temporal stability in school timing across years, allowing us to quantify how the term starts consistently amplifies transmission through child mixing, consistent with prior work linking school holidays to delays in epidemic peaks (*8*).

##### Built Environment

Ventilation with outdoor air can modulate influenza transmission by removing and diluting infectious virus in indoor air. We hypothesized that regional variation in building characteristics, including building age and shared ventilation systems, shapes the built-environmental context for transmission by affecting how much infectious virus accumulates indoors. Continuous, time-varying metrics (usage of mechanical heating/cooling, fan usage) further isolate specific indoor transmission mechanisms from these broader geographic gradients.

- Mechanical heating and cooling, derived from smart thermostat data (*57*), measures the proportion of time climate control (air conditioning or heating) is used. Heating and cooling can influence transmission in opposing ways: active use may reduce natural ventilation by keeping windows and doors closed, while air passing through the system is also diluted and filtered (*26–28*). Directed airflow from these systems can further carry infectious droplets and aerosols across a room (***?***).
- Complementarily, fan usage proportion, also from smart thermostat data, served as a proxy for mechanical air handling independent of heating and cooling demand (though the fan also runs during active heating or cooling calls). The same mechanisms, reduced ventilation, dilution, filtration, and directed airflow, plausibly apply. Geographic variation in fan usage independent of heating or cooling demand may reflect building codes requiring mechanical ventilation in tightly sealed, colder-climate housing.
- Housing vintage data from the U.S. Census Bureau (*51*) were used to quantify the proportion of homes built between 1950 and 2009. We chose this metric because these homes likely have tighter and lower infiltration rates compared to older buildings (pre-1950), which were constructed in an era of fewer concerns about energy conservation, and newer buildings (post- 2009), which were subject to updated mechanical ventilation standards introduced around 2010 (*58, 59*). This housing vintage may therefore represent homes with reduced natural air exchange but without the compensatory mechanical ventilation requirements of more recent construction.
- Multi-family housing prevalence, also from Census data (*51*), was included as a potential factor influencing transmission risk. Multi-family buildings typically have lower infiltration rates than single-family homes (*59*), potentially creating more stagnant air conditions that favor pathogen accumulation. Additionally, these buildings are associated with higher occupant density per building, which may influence transmission dynamics. These factors may create indoor environments that differ from single-family homes in ways that could influence aerosol pathogen transmission dynamics.
- To account for heterogeneity in housing across U.S. counties, we included the CDC’s Social Vulnerability Index (SVI) (*60*). Specifically, we used Theme 4 (“Housing and Transportation”), which captures structural and transportation-related vulnerabilities. This includes factors such as residence in multi-unit or mobile homes, household crowding, living in group quarters, and lack of vehicle access.

Given the heterogeneous spatial coverage and resolution across our eleven data sources, we implemented a comprehensive spatial imputation framework to ensure robust county-level estimates. Missing values for counties were estimated as the average of all directly adjacent counties with available data. All datasets are described in detail in the Supplementary Text.

##### Prior Immunity

To account for additional sources of heterogeneity in transmission dynamics, we incorporated imputed season and age specific weekly vaccination coverage at the county level from (*61, 62*), capturing heterogeneity in population immunity rather than assuming uniform susceptibility. We also included past county-level influenza prevalence, derived from our claims data, as a proxy for population immunity from prior infection.

#### Statistical Analysis of Influenza Seasonal Dynamics

To identify the factors associated with influenza seasonality patterns, we developed generalized linear models analyzing both summer incidence levels and epidemic onset timing across U.S. counties. For each target outcome, candidate predictors across regional mixing, social mixing, and built environment categories were entered simultaneously into a single multivariable GLM specification alongside season fixed effects. Prior to model estimation, pairwise multicollinearity diagnostics using variance inflation factors (VIF) were conducted, and highly correlated predictors (VIF > 5) were excluded to prevent variance inflation. For summer incidence, we tested competing hypotheses regarding viral circulation maintenance during typically low-transmission periods, using log-transformed normalized summer incidence (weeks 22–32) as the dependent variable, as defined above. Key explanatory variables included the three categories of predictors: regional mixing, social mixing, and built environment. Time-varying predictors (e.g., indoor activity) were summarized as mean over weeks 22-32.

For onset timing analysis, we computed the regression with the same predictors used for summer incidence (except for household occupancy, which was excluded due to collinearity), adding summer incidence and school calendars as predictors. The dependent variable was the week of epidemic onset. We also included spatial connectivity to early-onset counties to capture geographic propagation patterns. All variables were averaged over weeks 29-34 to capture conditions during the epidemic seeding phase, spanning the weeks immediately preceding onset (29–33) through the earliest observed onset (34). This window provides a temporally coherent summary of the transition from summer baseline to epidemic conditions, which we hypothesized would be most informative for predicting onset timing. All predictor variables were z-normalized to enable direct comparison of effect sizes across different measurement scales. Multicollinearity diagnostics using variance inflation factors identified and excluded highly correlated predictors (VIF > 5). To account for spatial and temporal dependence in the data, standard errors were computed using Driscoll–Kraay heteroskedasticity- and autocorrelation-consistent estimators, which are robust to general forms of cross-sectional (spatial) correlation and serial correlation across time. Final models employed Gaussian regression with identity link functions. Both analyses covered influenza seasons 2016/2017 through 2019/2020, excluding post-2020 seasons due to SARS-CoV-2 pandemic-induced disruptions that fundamentally altered influenza epidemiological patterns.

#### Mechanistic Transmission Modeling

To mechanistically understand what shapes spatial and temporal heterogeneity in influenza seasonality, we developed a stochastic, age-structured metapopulation transmission model at the US county level. The model implements SEIR (Susceptible-Exposed-Infectious-Recovered) compartmental dynamics with influenza-specific epidemiological parameters. We stratified the population into two age classes: 5-17, 18-65. We focused on these age groups because they drive the vast majority of population-level community transmission and routine social mixing, whereas high-risk groups (< 5 and > 65) predominantly reflect severe clinical outcomes rather than baseline transmission dynamics. The model framework builds upon established approaches for spatially-coupled disease dynamics (*54*), incorporating three transmission pathways: (i) local transmission among residents within their home county, (ii) importation from infected visitors arriving from other counties, and (iii) exportation through residents who acquire infection during travel to other counties and return home. Transmission rates were modulated by county-specific contact variation (*55*) and weekly age-specific contact rates derived from established mixing matrices (*8*). The matrices were modified to vary based on school calendar status and holiday calendars, with elevated mixing among school-age children during term time.

To capture the population immune landscape, we integrated vaccination coverage by week (Fig. S13) into the model but not our proxy for prior infection immunity (based on the results of our onset timing statistical analysis).

The baseline transmission parameter, *β*_0_, was calibrated to reproduce observed national incidence from week 31 through week 6 of the following year (27 weeks), spanning the full epidemic season from late summer through winter peak. The model is seeded at week 31 using observed domestic case counts for each county, adjusted and normalized for underreporting in the claims data (Supplementary Text), so initial conditions are empirically anchored rather than estimated as free parameters. Simulation begins at week 32, two weeks before the earliest observed onset (week 34), allowing transmission dynamics to develop from these seeded conditions rather than imposing onset as an initial condition. Because the metapopulation is connected across counties, this seeding scheme permits multiple seeding events, bidirectional spread, and simultaneous emergence at several locations as outcomes of the model.

We simulate each county as an isolated population (no mobility scenario). We then systematically tested the contribution of different mechanistic components: i) spatial connectivity and contact variation; and ii) connectivity, contact variation with school calendar effects. We then integrate local transmissibility modulation based on crowding and poor ventilation using a statistical approach (described below). We evaluated the relative importance of each set of factors in predicting epidemic onset timing. Detailed model equations, calibration procedures, and sensitivity analyses are provided in the Supplementary Text.

##### Empirical Integration of Local Transmissibility

While it is well established how to mechanistically integrate mobility and contact patterns into transmission models, we lacked a framework for incorporating the effects of built environment factors on transmission. To quantify and integrate these transmission factors, we developed a residual transmission enhancement framework that identifies and parameterizes their contributions. We first executed the baseline mechanistic transmission model incorporating vaccination, contact variation, school-driven contact pattern changes, intercounty mobility networks, and observed summer incidence seeding across all U.S. counties. From the simulated county-level incidence trajectories, we calculated instantaneous growth rates using the same methodology applied to observed data (above), and computed the transmissibility, *β_model_* (*t*), from these simulated growth rates using:

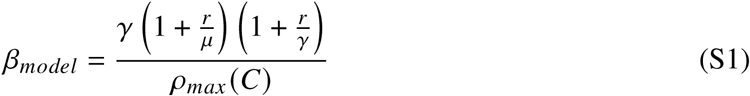

where *r* is the instantaneous growth rate, *γ* is the recovery rate (inverse of the mean infectious period), *μ* is the incubation rate (inverse of the mean latent period, corresponding to the rate at which exposed individuals become infectious), and *ρ_max_* (*C*) is the maximum eigenvalue of the contact matrix.

Parallel analysis of observed national influenza incidence yielded the empirical transmissibility, *β_obs_* (*t*), required to reproduce observed epidemic patterns using the same formulation. The residual transmissibility Δ*β*(*t*) = *β_obs_* (*t*) − *β_model_* (*t*) quantifies the transmission not accounted for by vaccination, contact, school, mobility, and initial seeding effects alone.

To empirically characterize these residual effects, we developed a generalized additive model (GAM) with Δ*β*(*t*) as the dependent variable. Predictors included variables for which mechanistic transmission relationships have not been established: indoor-outdoor activity ratios, mechanical heating/cooling/fan usage, household size, building age and occupancy characteristics, multi-unit housing prevalence, and social vulnerability, along with a smooth term for weeks since epidemic onset to flexibly capture residual temporal structure not explained by the parametric predictors This regression framework enabled the decomposition of residual transmission variability into quantifiable components, generating predicted transmission enhancement values Δ*β*(*t*) based on observed built environment conditions. We then integrated these predicted residual effects into our transmission model as a multiplicative factor applied to the force of infection. The final integrated model thus captures both explicitly parameterized behavioral mechanisms (school mixing, spatial connectivity) and empirically-derived built environment predictors that collectively determine seasonal influenza transmission dynamics.

#### Evaluation of Vaccination Strategies

Using the parameterized metapopulation model as a baseline representing status quo US influenza vaccination timing and coverage, we simulated two alternative vaccination strategies: a uniform national rollout, shifted 4 or 8 weeks earlier than the observed rollout, and a geographically targeted, onset-based rollout, implemented locally 4 or 8 weeks before each county’s predicted epidemic onset. For the onset-based strategy, we defined local onset timing using both observed onset weeks as well as predicted onset weeks based on our model (as observed onset timing would be unavailable in real time). Because earlier vaccination allows more time for immunity to wane before local epidemics arrive (*32*), we evaluated each timing strategy with and without waning immunity. Waning vaccine effectiveness was implemented using empirical estimates from a within-season case-control study (*31*), with vaccine effectiveness declining from 59% at 14-29 days post-vaccination to 52% at 30-59 days, 39% at 60-89 days, and 34% at 90+ days. For each scenario, we compared total cases prevented relative to baseline and assessed the impact of each strategy on the spatial progression of the epidemic using rank correlations between baseline and scenario onset weeks across counties.

### Supplementary Text

#### Data Summary

Our analysis focuses on the 2018-2020 period to maximize dataset overlap while excluding the early COVID-19 diffusion period (2020–2021), when non-pharmaceutical interventions disrupted regular activity patterns and influenza seasonality. All datasets span this analysis window 2018-2020 except school calendar data, which we collected for the 2023-2024 academic year under the assumption that school schedules remain consistent across years.

##### Medical Claims Data

To capture fine-scale temporal and spatial patterns of influenza activity across the United States, we utilized high-volume medical insurance claims data spanning 2016-2024, derived from a commercial healthcare clearinghouse that processes transactions between healthcare providers and insurance payers. These de-identified claims data, representing approximately 9.5 billion claims from an average of more than 133 million patients annually, provide county-day resolution time series of confirmed influenza cases based on International Classification of Diseases 10th revision (ICD-10) codes, offering substantial advantages over traditional surveillance systems in terms of geographic granularity and temporal resolution. The dataset captures approximately 40% of county populations, on average, with good representation across both private and public insurance sources, enabling robust detection of local epidemic dynamics that might be obscured in coarser surveillance data. We also extracted respiratory syncytial virus (RSV) cases from the same claims database to compare respiratory disease seasonality patterns. Cases are corrected to count only the earliest infection per patient per flu season, ensuring accurate representation of disease incidence rather than healthcare-seeking behavior.

To account for variations in healthcare-seeking behavior and insurance coverage across counties and periods, we applied a two-step adjustment and normalization procedure. First, we corrected for temporal fluctuations in healthcare-seeking/reporting volume within each county by scaling weekly confirmed case counts by a county-specific weekly measurement factor, defined as that county’s weekly all-cause visit volume relative to that same county’s own average all-cause volume for the year. This factor is computed independently for each county using only that county’s own data, correcting for within-county temporal reporting fluctuations (e.g., holidays, local careseeking patterns) without requiring comparable coverage across counties. Second, we normalized these adjusted case counts by that same county’s total all-cause visit volume, expressing incidence relative to each county’s own healthcare-seeking population rather than as an absolute count. It is this second step that renders the resulting metric comparable across counties with differing absolute levels of data coverage, since incidence is expressed per unit of each county’s own measured healthcare activity rather than assuming similar raw coverage nationally. A geographic assessment of coverage across US counties, showing no strong systematic heterogeneity, along with additional details on this data source, are available in (*63*).

**Table S1:** Multi-source data integration. Complementary datasets to quantify influenza and RSV seasonality and its predictors. POI denotes point of interest.

| Data Source | Key Variables | Resolution | Period | Sample Size | Coverage |
| --- | --- | --- | --- | --- | --- |
| Medical Claims Data | Influenza cases, RSV cases | Individual-day | 2016–2024 | 9.5B claims | 145M patients/yr |
| School Calendar Data | School start dates, holiday periods | District-day | 2023–2024 | 8,784 districts | – |
| Contact Data | Non-household interactions | County | Pre-2020 (est.) | 3,100 counties | 10.7M survey responses |
| Mobility Data<br>(SafeGraph/Advan) | County-to-county movement flows | POI-week | 2018–2021 | 4.6M POIs | 45M+ devices |
|  | Indoor vs. outdoor visit propensity | POI-week | 2018–2021 | 4.6M POIs | 45M+ devices |
| Air Traffic Data | Passenger volumes from South America | Airport-month | 2016–2020 | 3,944 airports | ~994M/yr |
| WHO FluNet | Influenza circulation in South America | Country-week | 2016–2020 | – | – |
| Smart Thermostat<br>(ecobee) | Mechanical heating, cooling, fan usage | Building-second | 2017–2024 | 230,122 thermostats | 230,122 buildings |
| Census Data (ACS)<sup>1</sup> | Household size, occupancy, houses built 1950–2009, multi-unit housing (%) | County-year | 2016–2023 | 820–3,143 counties | – |

These data are natively recorded at the individual patient-encounter level; patient encounters were geolocated and spatially aggregated to the county level based on the patient’s five-digit residential ZIP code.

##### School Calendar Data

To incorporate the impact of school schedules on age-specific mixing patterns and influenza transmission dynamics, we utilized a publicly available dataset of school calendar data at the US school district level from public school districts across the United States for the 2023-2024 academic year (https://publicholidays.com/school-holidays/, last accessed in Dec 2024). We focused specifically on school start dates and winter holidays from this dataset, as these mark the transition from summer break to increased social mixing among school-age children with a break for the holiday period. We assumed that the spatial pattern of school start dates remains stable across years, allowing us to apply the 2023-2024 calendar patterns to our study period (2106–2020). We used age-specific contact rates based on school calendar status for two age classes, 5-19 and 19-65, with school start dates triggering elevated contact rates for the school-age population to reflect increased mixing in classroom and school environments (*8*). These time-varying, age-structured contact patterns have been estimated in the POLYMOD study (*64*).

Data were natively collected at the public school district level; district-level start dates were spatially aggregated to the county level by mapping each school district to its corresponding county (assigning the population-weighted modal start date when a county contained multiple districts).

##### Mobility Data

###### Inter-county Connectivity Network

To understand domestic importations and geographical patterns of influenza onset timing, intercounty connectivity serves as a critical mechanism for viral spread between regions with different seasonal dynamics. We utilized high-resolution mobility data from SafeGraph (now Advan Patterns) spanning 2019-2021, which captures movement patterns between U.S. counties based on mobile device location data from over 45 million devices. We aggregated daily visits between census block groups to the county level and calculated intercounty connectivity by normalizing visits from origin to destination counties. Given our previous findings that demonstrate that inter-county connectivity networks remain temporally stable (*54*), we used a weekly static connectivity matrix averaged in 2019 to represent baseline mobility patterns between counties. These connectivity networks enable us to quantify how human movement facilitates the geographical spread of influenza across different regions, particularly from counties with early onset timing or high summer incidence to more susceptible areas.

This dataset was natively collected at the Point of Interest (POI) level; visit counts were spatially aggregated to the county level by assigning each POI to its containing county using latitude/longitude coordinates.

###### Indoor Activity Index

To quantify increased transmissibility rates in locations where populations spend more time indoors, we utilized the indoor activity seasonality metric previously computed by (*10*). This metric, derived from SafeGraph mobility data, quantifies seasonal variation in indoor versus outdoor human activity at the county-week level by classifying approximately 4.6 million points of interest using North American Industry Classification System codes as primarily indoor (90% of locations, including schools, hospitals, stores) or outdoor (6.5% of locations, including parks, outdoor recreation areas). The indoor activity metric captures the propensity for visits to be to indoor locations relative to outdoor locations, normalized by maximum visit counts and mean-centered for comparability across counties. These indoor activity patterns provide a quantitative measure of seasonal exposure risk for respiratory diseases like influenza, as increased indoor activity during certain periods and in specific places may enhance transmission potential through prolonged co-location in enclosed environments. When coupled with ventilation rate measurements, this metric enables us to identify locations and periods with heightened transmission risk due to behavioral patterns that increase indoor crowding and reduce air quality.

**Figure S1:**
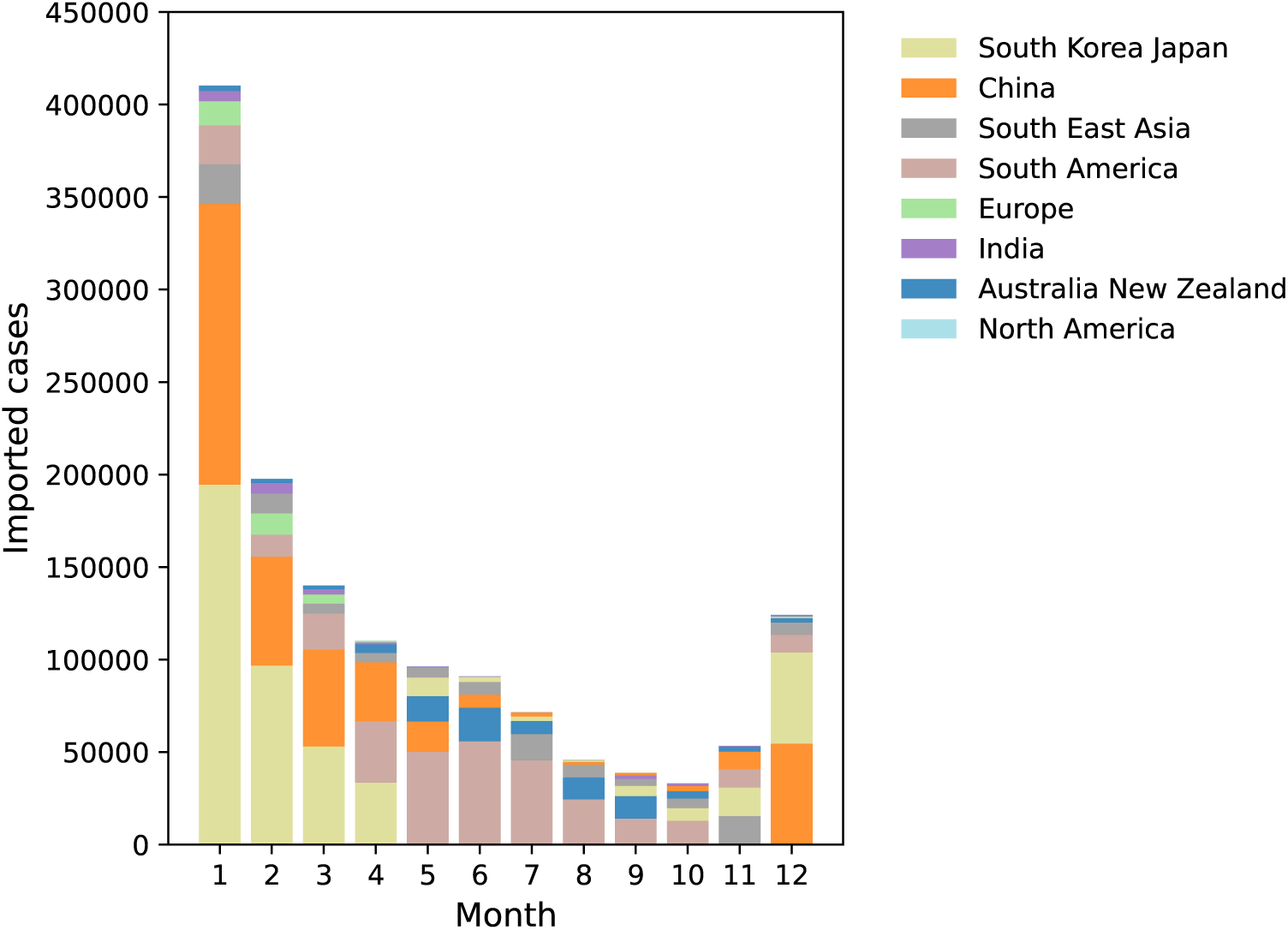
Monthly number of imported cases. Stacked bar chart showing the mean number of imported cases per month aggregated across years from 2016 to 2020, stratified by geographic region of origin. Each bar represents the total monthly imported cases, with colors indicating the relative contribution from different regions.

##### Air Traffic and WHO FluNet Data

To quantify a proxy measure for international importation of influenza from southern America during the US summer months, we integrated World Health Organization (WHO) influenza surveillance data with U.S. air passenger traffic data from the Bureau of Transportation Statistics (BTS). WHO surveillance data provided comprehensive daily country-level influenza information, including cases across different strain types, test positivity rates, and temporal patterns of circulation during their winter flu seasons (May-September) (*65*). BTS air traffic data provided monthly passenger volumes from each country to US airports (https://www.transtats.bts.gov/airports.asp). We select data from 2016 to 2019 from both datasets and mapped to a weekly scale. We estimated infection rates among arriving passengers by deriving time-varying positivity rates for each origin country. For each country, we identified the peak incidence period (maximum total influenza cases) and extracted the corresponding test positivity rate during that peak. We then scaled positivity rates for all other periods proportionally to the observed incidence levels, using the peak incidence and peak positivity rate as reference points. This approach generated realistic infection probabilities that varied temporally with disease circulation intensity in each origin country and minimized confounding by country-specific surveillance capacity.

We applied these country-specific, time-varying infection rates to weekly passenger flows to estimate the number of potentially infected passengers arriving at U.S. destinations every week. To account for the spatial distribution of international arrivals beyond the US county of airports, we allocated incoming passengers from each origin country to destination counties and their neighboring counties based on population weights, recognizing that international travelers may disperse throughout surrounding regions. This approach generated county-level estimates of weekly international importation pressure from southern America sources, providing a quantitative measure of how global connectivity patterns during opposing seasonal cycles might contribute to the maintenance of summer influenza circulation and influence subsequent epidemic onset timing across different US regions.

Our estimate for summer international importations in the US, mainly coming from South America, is shown in Fig S1, and in agreement with (*34*).

##### Smart Thermostat and Weather Data

To quantify indoor environmental conditions and ventilation patterns that can influence the transmission of respiratory diseases, we used data from *ecobee* smart thermostats over the period 2017-2024 (*57*). These devices provide high-resolution measurements of indoor temperature, indoor humidity, and mechanical heating/cooling/fan usage patterns from residential buildings across the United States. Each thermostat identifier was assigned to a US county based on state and city information using OpenCage’s Geocoding API. For each thermostat, we determined the most frequently appearing county within each year to account for potential device relocations. This data represents 0.1 % of buildings (primarily residential) across the US. We restricted our analysis to counties with at least 10 thermostats sampled in the dataset to ensure robust representation of local indoor conditions. We processed the raw telemetry data, which consists of 5-minute interval measurements, through several aggregation steps. For each thermostat, we determined whether occupancy or heating/cooling/fan activity occurred during each measurement interval. We then aggregated these individual thermostat measurements to the county-week scale to measure the average proportion of time that heating/cooling/fans were active and buildings were occupied in a given county-week. While smart thermostat ownership is likely to skew toward wealthier and urban areas, 89% of US households use air conditioning (80% among the lowest-income groups) (*66*), suggesting similar behavioral cooling responses across populations. Housing differences, including building age and structural vulnerabilities affecting ventilation, are captured in other variables of the analysis.

In the analysis, High use of mechanical heating/cooling indicates closed building systems with active climate control (air conditioning or heating), suggesting poor natural ventilation and reduced air exchange with the outdoor environment. Additionally, high fan usage (outside of mechanical heating/cooling) serves as a measure of recirculated air. These combined metrics provided county-level indicators of building ventilation patterns that could influence aerosol transmission dynamics, particularly during periods when populations spend increased time in poorly ventilated indoor environments.

#### Onset week detection

Epidemic onset timing was estimated from growth rate dynamics. We calculated the instantaneous growth rate of influenza incidence and identified candidate onset periods when the growth rate exceeded *r* > 0.1 for at least three consecutive weeks. To reduce fluctuations in growth rate calculations, we set case counts below 8 to zero, as very low case numbers produce highly volatile growth rate estimates that frequently fluctuate above the threshold without representing true epidemic growth. When multiple time windows met these criteria—for example, due to transient early-season increases followed by decline before sustained epidemic growth—the onset detection algorithm selected the most epidemiologically relevant window by weighting three factors: (1) the duration of consecutive days above the growth rate threshold, (2) the absolute incidence level during the candidate window, and (3) the mean growth rate magnitude within that window. This multi-criteria approach ensures that detected onsets correspond to sustained epidemic signals rather than statistical artifacts from low case counts or brief fluctuations. We estimated the onset week for each county across all influenza seasons in our study period.

#### Spatiotemporal patterns of influenza transmission in the United States

**Figure S2:**
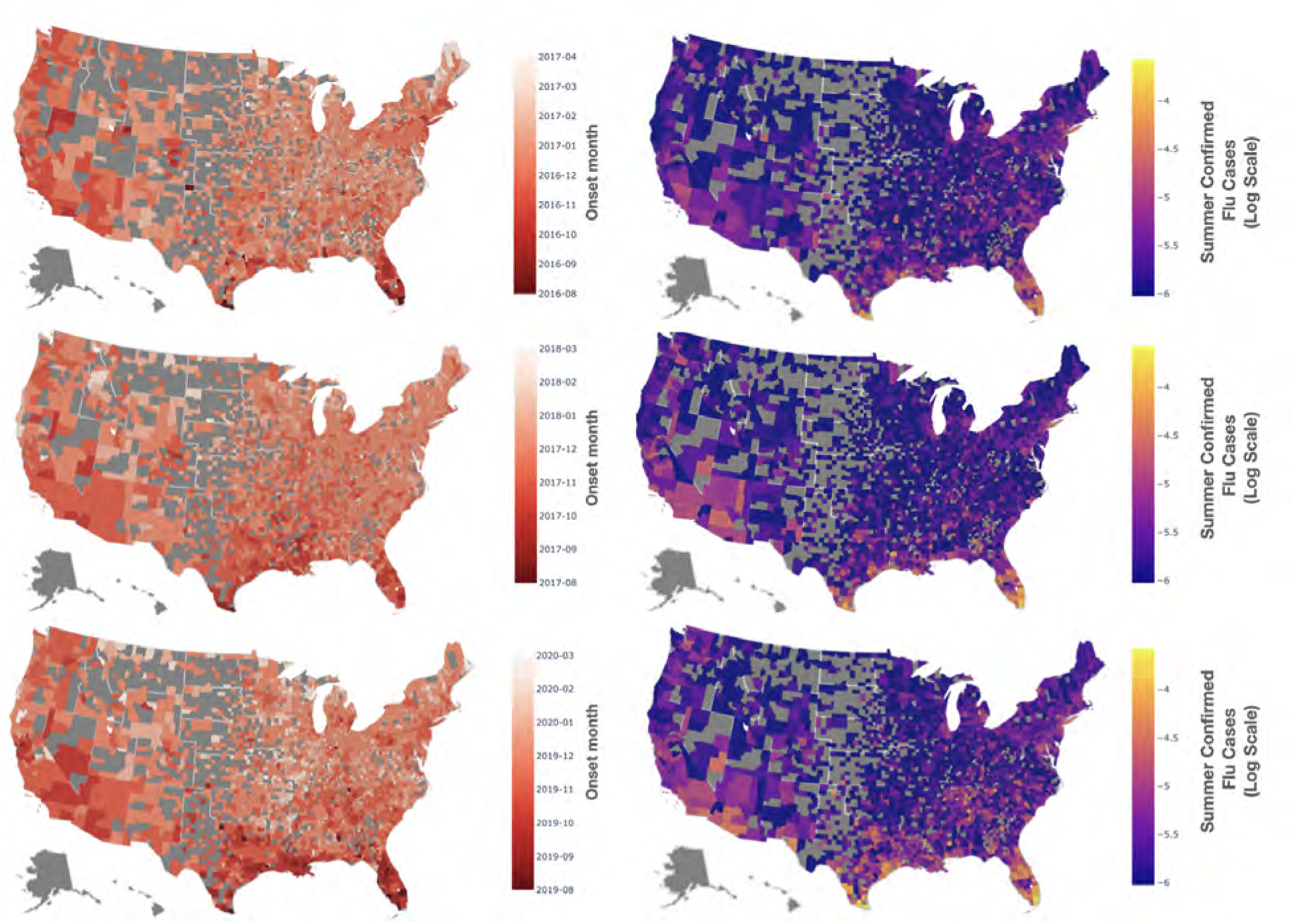
Spatiotemporal patterns of influenza transmission across multiple epidemic seasons. County-level maps showing epidemic onset timing and summer influenza incidence (log scale) for three additional seasons: 2016-2017, 2017-2018, and 2019-2020. Consistent patterns emerge across all seasons, with persistent summer circulation concentrated in southern US counties and epidemic onset following the characteristic south-to-north progression. These complementary seasons demonstrate the robustness of the spatiotemporal dynamics observed in the main analysis, confirming that the mechanistic predictors of summer persistence and epidemic timing operate consistently across different influenza seasons and strain compositions.

#### Post COVID-19 Influenza Seasonality analysis

**Figure S3:**
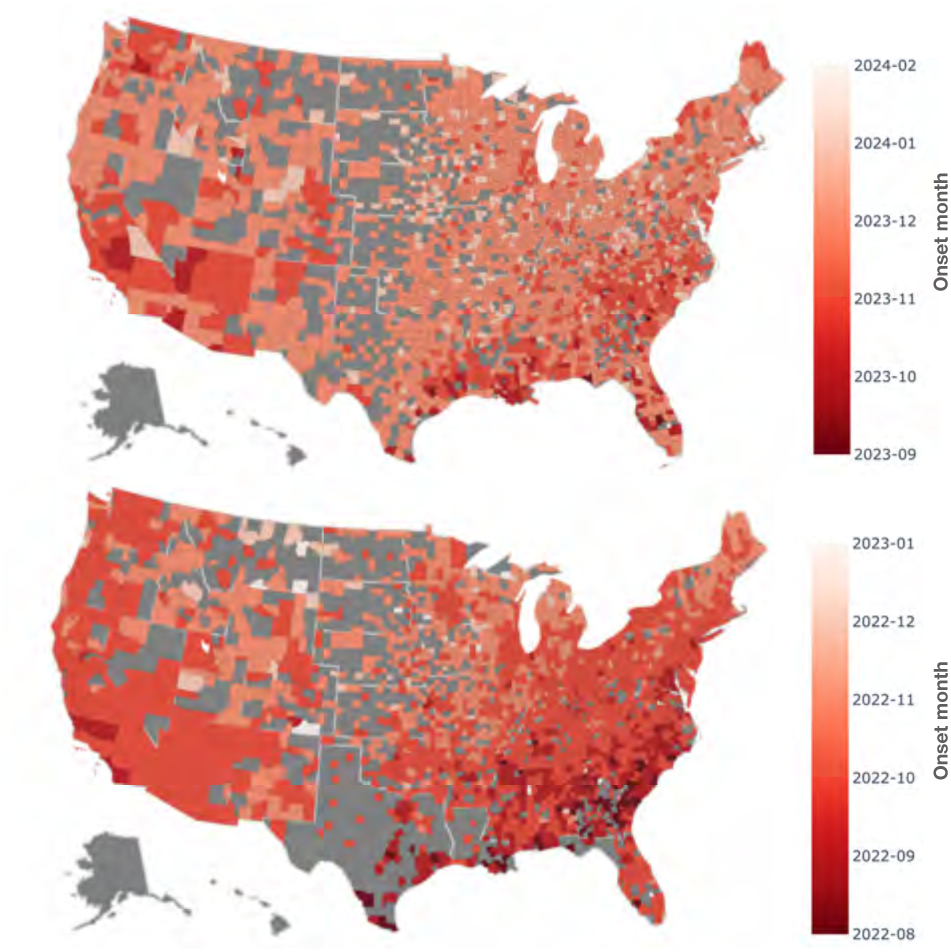
Geographic Heterogeneity of onset timing during post-COVID periods. Maps show the timing of onset across US counties for 2022/2023 (bottom) and 2023/2024 (top) seasons. Earlier onset is indicated by darker colors, while lighter colors represent later onset timing.

#### Characterization and Distinction of Reservoir vs. Bridging Hub Counties

To address the mechanism of seasonal persistence versus early epidemic importation, we partitioned counties based on their summer persistence and onset timing into two distinct operational categories:

1. **Summer Reservoir Counties:** Counties that sustain continuous baseline transmission through the epidemiological low season (weeks 22-32). These counties are operationalized as maintaining a median weekly incidence rate exceeding the national summer baseline during this low-season window.
2. **Bridging Hub Counties:** Urban counties characterized by high inter-county connectivity that receive early viral importation, exhibiting epidemic onset in the earliest quartile (≤ *Q*_25_) for their respective latitude band but do not sustain summer circulation above the national threshold (i.e., non-reservoirs).

Summer reservoir counties cluster predominantly at lower latitudes (< 30^◦^N) across all analyzed seasons (Figure S4). In contrast, major northern metropolitan centers such as New York County (NY) and Cook County (Chicago, IL) remain consistently below the national summer baseline during weeks 22-32 in every season. This empirical distinction confirms that highly connected northern urban centers do not act as local summer reservoirs. Instead, their role is that of a bridging hub: rather than seeding the autumn wave through persistent summer transmission, they receive imported viral activity early in the season and rapidly redistribute it across surrounding regional networks.

**Figure S4:**
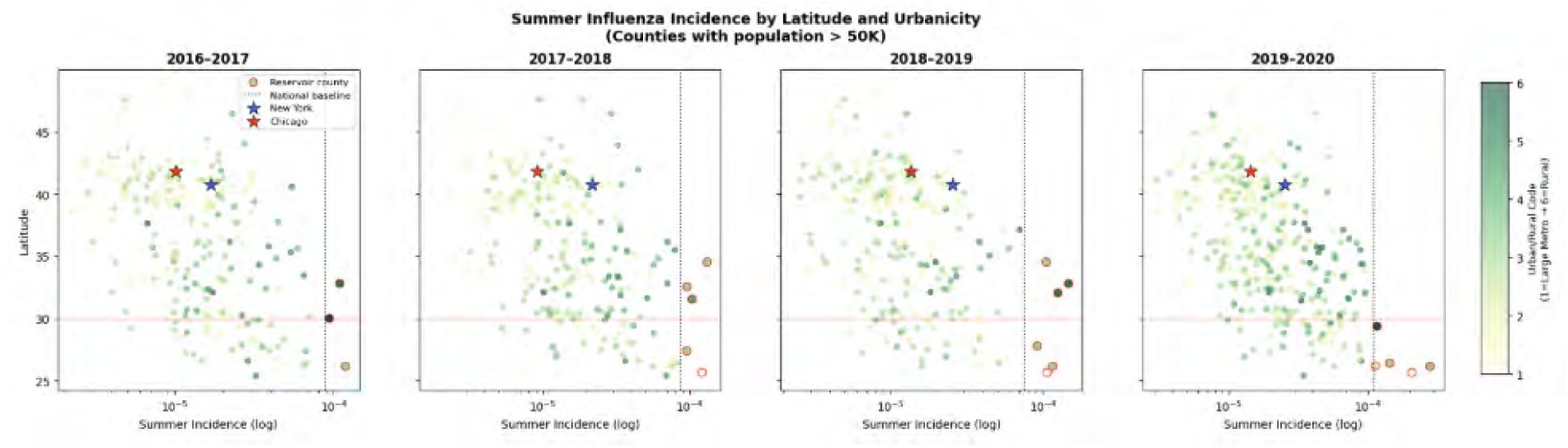
Median Summer Influenza Incidence vs. Latitude across Four Epidemic Seasons. Scatter plots display median weekly incidence during the inter-epidemic summer period (weeks 22-32) as a function of county latitude across four seasons. The vertical dotted line indicates the national summer baseline threshold. Red-bordered points denote identified summer reservoir counties exceeding this baseline. Major northern metropolitan hubs, including New York County (blue star) and Cook County/Chicago (red star), lie to the left of the national baseline across all seasons, confirming that they do not sustain summer-season circulation at reservoir levels.

To evaluate the relative contributions of geographic position and urban connectivity to epidemic timing, we analyzed county-level onset week across four pre-pandemic seasons (2106–2020) as a function of latitude and the National Center for Health Statistics (NCHS) Urban-Rural Classification scheme (codes 1-6, where 1 represents Large Central Metro and 6 represents Rural).

We fit an Ordinary Least Squares (OLS) regression model specifying onset week (*Y_i_*_,*t*_) for county *i* in season *t*:

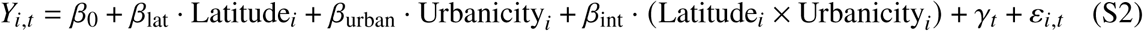

where continuous predictors were *z*-standardized across all *n* = 8, 752 county-season observations to enable direct comparison of effect sizes (*β*), and *γ_t_* represents season fixed effects.

**Table S2:** Multivariate OLS regression predicting epidemic onset week timing (*n* = 8, 752). Predictors were *z*-standardized.

| Predictor Variable | Coeff ($\beta$) | Std. Error (SE) | $t$-statistic | $p$-value |
| --- | --- | --- | --- | --- |
| Urbanicity (NCHS Gradient) | −0.96 | 0.04 | −24.00 | < 0.001 |
| Latitude (°N) | +0.94 | 0.04 | +23.50 | < 0.001 |
| Latitude × Urbanicity Interaction | +0.03 | 0.04 | +0.75 | 0.404 |
| $R^2 = 0.30$ | | | | |

Urbanicity and latitude yielded nearly identical standardized effect sizes (*β*_urban_ = −0.96, *β*_lat_ = 0.94, both *p* < 0.001), indicating that the urban/rural gradient contributes as much to explaining onset timing as geographic latitude itself. When treating urban/rural status as a continuous gradient, each standard deviation increase toward greater rurality was associated with 1.43 additional weeks of delay in epidemic onset (*β* = 1.43, SE = 0.04, *p* < 0.001, *R*^2^ = 0.18). The interaction between latitude and urbanicity was not statistically significant (*p* = 0.404), demonstrating that this urban onset advantage is consistent across all latitude belts.

**Figure S5:**
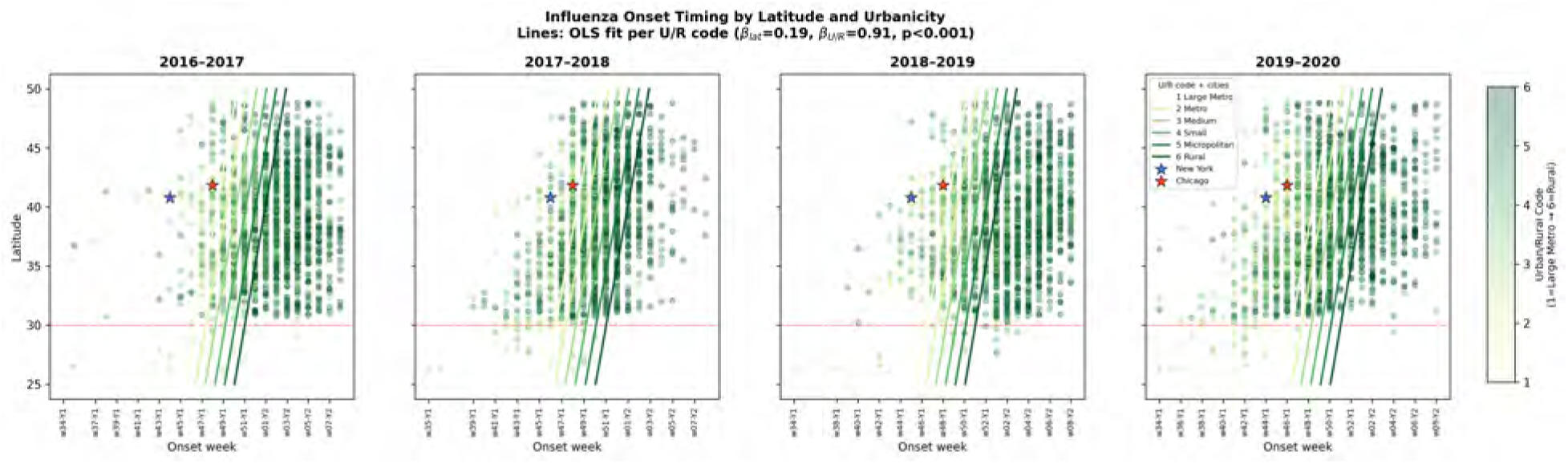
Influenza Onset Timing by Latitude and Urbanicity across Four Epidemic Seasons (2106–2020). Scatter plots show county-level epidemic onset week versus latitude for each season (2016–2017 through 2019–2020). Points are colored according to NCHS Urban/Rural Code (1 = Large Central Metro [light green] to 6 = Rural [dark green]). Blue-bordered circles highlight identified bridging hubs (urban, non-reservoir counties with onset ≤ *Q*_25_ for their latitude band). Key metropolitan centers—New York County (blue star) and Cook County/Chicago (red star)—are explicitly highlighted among the earliest onsets at their latitude. The red horizontal dashed line indicates 30^◦^N latitude. Overlaid solid lines display fitted OLS regression slopes per urban-rural code (*β*_U/R_ = 0.91, *p* < 0.001), illustrating the systematic urban onset advantage across all latitudes.

##### Stability of Regression Coefficients Across Seasons for Summer Incidence and Onset Timing

To evaluate the temporal consistency and robustness of our empirical findings across individual epidemic cycles, we performed season-stratified multivariable GLM regressions for each prepandemic season from 2016 to 2019 (Figure S6). Overall, structural and demographic predictors governing baseline summer transmission intensity demonstrated interseasonal stability. Variables reflecting high-density built environments and housing vulnerabilities—such as multi-family housing prevalence, population density, housing vintage (built 1950-2009), and contact heterogeneity—maintained strong, positive associations with summer incidence across all four seasons. Similarly, regional seeding via international infected air passenger arrivals from South America and local social mixing metrics (contact, household size and indoor activity) consistently yielded positive point estimates across individual years. In contrast, mechanical recirculation, measured by fan usage, maintained a stable, negative association across seasons.

Conversely, predictors characterized by transient environmental and meteorological fluctuations exhibited higher interseasonal volatility. Built environment variables [CHECK] showed wider confidence interval spreads and point estimate shifts across years, reflecting interannual variability. This contrast confirms that while the baseline spatial distribution and magnitude of summer viral persistence are anchored by stable, structural features of the built environment and human demographic organization, local temporal dynamics and onset timing remain sensitive to transient, season-specific conditions.

#### Modulating Role of School Schedules on Epidemic Onset Prediction

To evaluate the specific contribution of school calendar dynamics to epidemic timing in counties with high summer incidence (top 10th percentile), we compared prediction metrics between the baseline model—incorporating spatial connectivity and contact heterogeneity—and the expanded model, which incorporates explicit county-level school reopening schedules and associated changes in age-specific contact mixing.

Incorporating school schedules provides a modest, incremental refinement in predicting epidemic onset timing rather than serving as a primary predictor of seasonality. Quantitatively, incorporating school dynamics reduces the Mean Absolute Error (MAE) of onset week predictions from 2.86 weeks to 2.68 weeks, with a notable reduction in extreme late-onset over-predictions, as shown in Figure S7.

**Figure S6:**
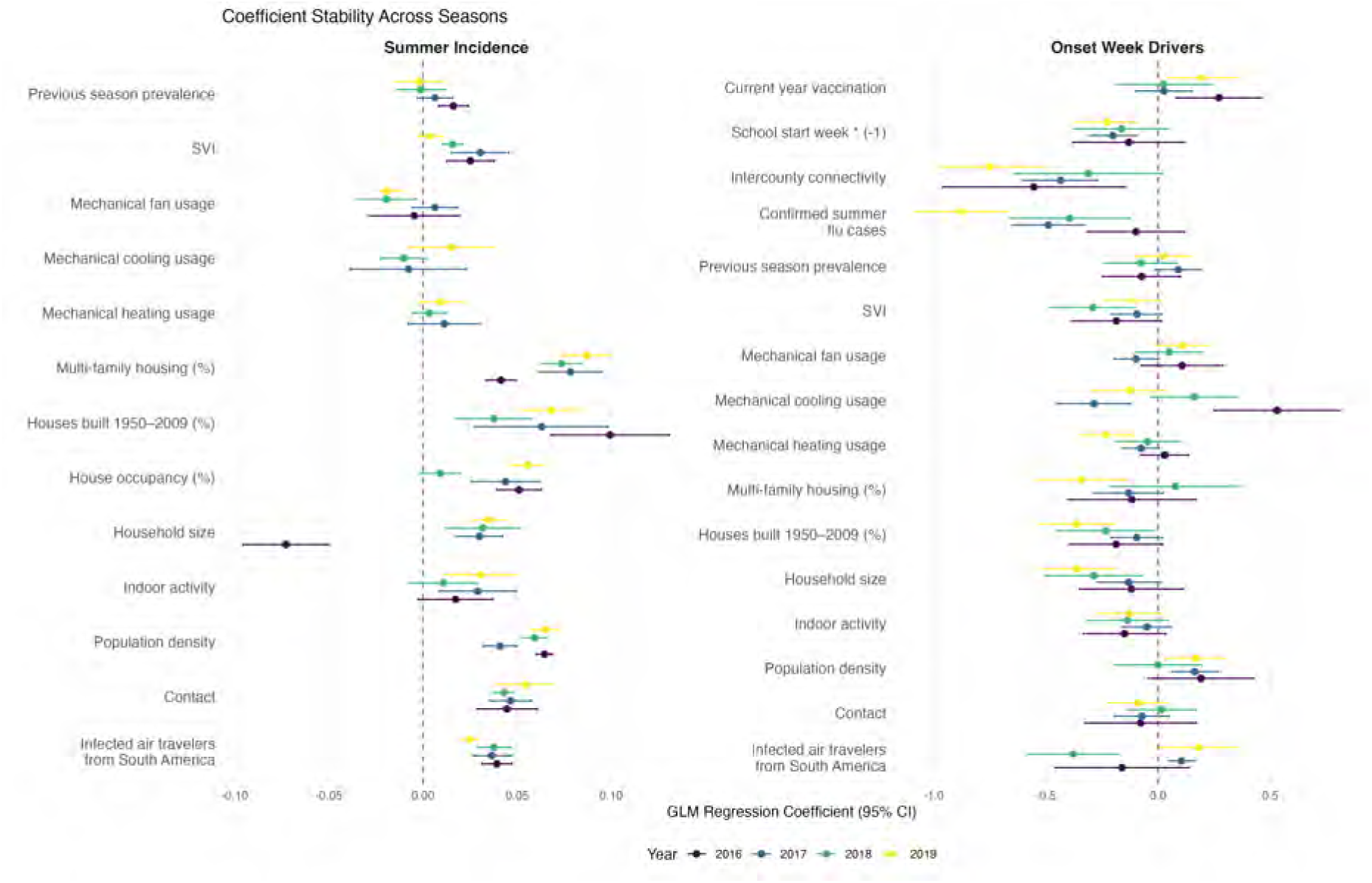
Interseasonal stability of multivariable regression coefficients for summer influenza and onset timing predictors (2106–2019). Standardized GLM regression coefficients and 95% confidence intervals estimated separately for each pre-pandemic influenza season: 2016 (purple), 2017 (blue), 2018 (green), and 2019 (yellow).

While rank-order correlation across counties remains uniformly high between models (Weighted Spearman correlation ≈ 0.93), the inclusion of school calendars improves absolute agreement with observed data, increasing the Weighted Concordance Correlation Coefficient (CCC) from 0.796 to 0.828. This metric alignment confirms that school reopening acts primarily as a temporal synchronizer: while intercounty connectivity remains the primary predictor of spatial dissemination, school schedules provide a localized secondary boost that accelerates transmission emergence in communities with established summer seeding.

**Figure S7:**
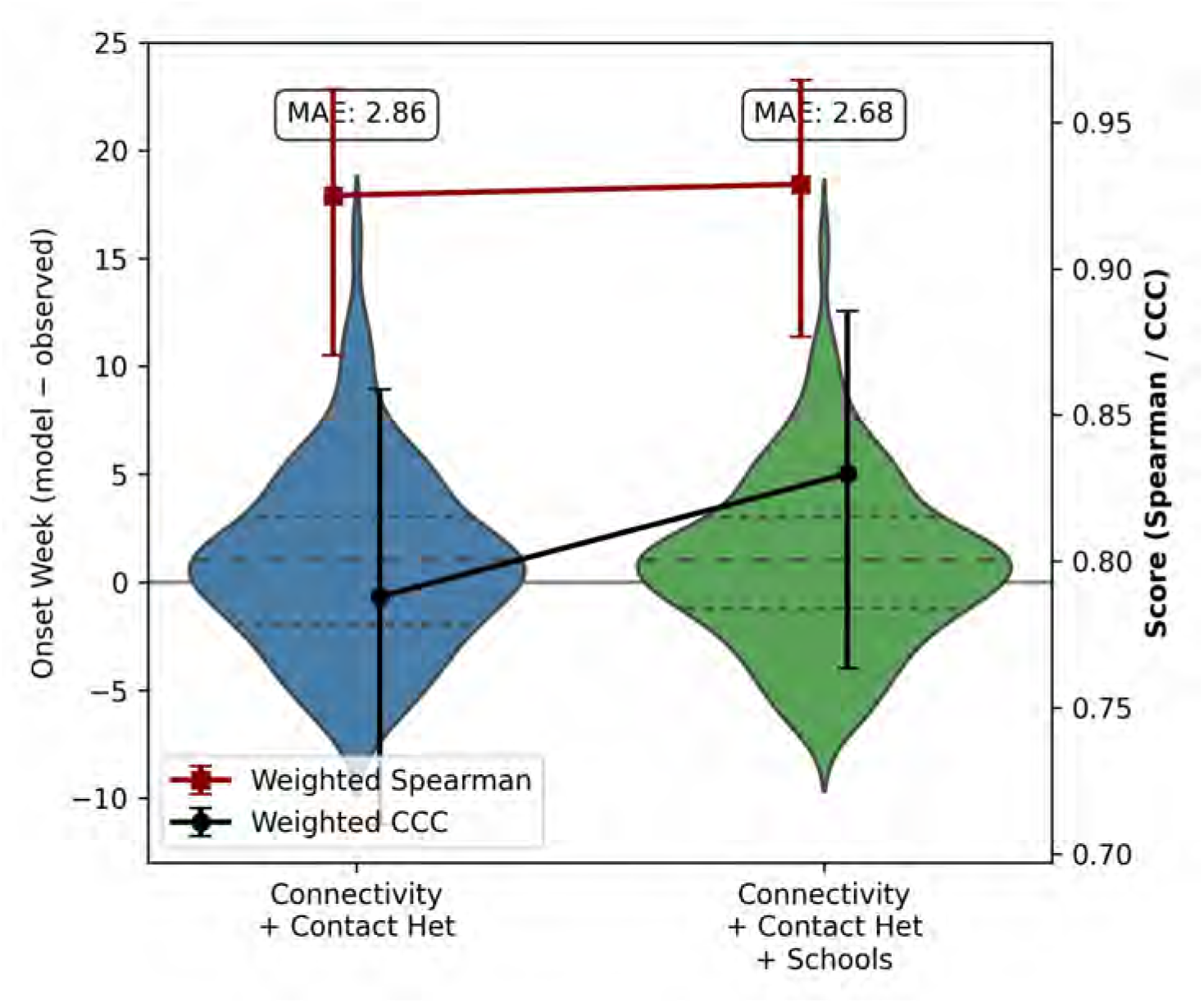
Impact of school calendar integration on epidemic onset prediction accuracy. Violin plots illustrate the distribution of prediction errors (simulated minus observed onset week) across U.S. counties for the model with connectivity and contact heterogeneity versus the model incorporating school schedules (+ Schools). Overlaid point estimates and error bars represent the Weighted Spearman correlation (red) and Weighted Concordance Correlation Coefficient (CCC, black) on the right axis. Text boxes display the overall Mean Absolute Error (MAE) in weeks for each model configuration.

#### Unexplained Variability in Onset Timing

To assess whether the global spatial autocorrelation in county-level residuals from the transmission model reflects a broad spatial process or a localized artifact, we computed Local Indicators of Spatial Association (LISA) using Queen contiguity weights. Global Moran’s I was moderate and statistically significant (I = 0.261, p = 0.001, 999 permutations), and this result was robust to using an alternative k-nearest-neighbor weighting scheme (I = 0.225, p = 0.001) and to excluding the most visually prominent cluster of counties in Michigan (I = 0.246, p = 0.001). Local decomposition (Figure S8), however, showed that only 12.2% of counties (198 of 1,622) formed part of a statistically significant local cluster (High-High or Low-Low, p < 0.05); the remaining 82.7% showed no significant local spatial association. Significant clustering was concentrated in a small number of discrete regional pockets, most notably counties in Michigan, Wisconsin, and Indiana.

**Figure S8:**
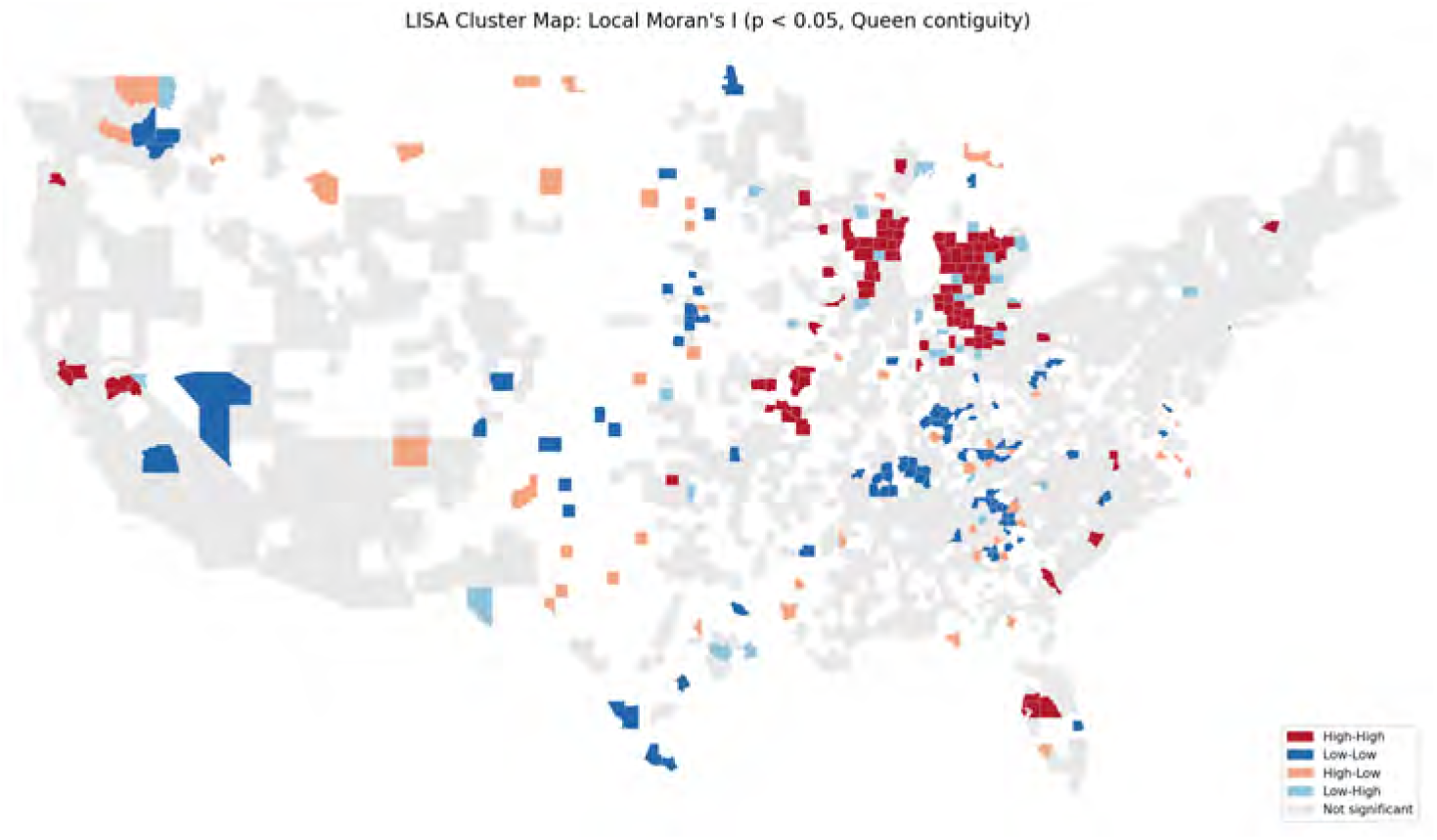
LISA cluster map of county-level residuals of onset times from the transmission model. Local Moran’s I clusters (Queen contiguity, 999 permutations, *p* < 0.05). Red indicates High-High clusters (positive residuals surrounded by positive residuals) and blue indicates Low-Low clusters (negative residuals surrounded by negative residuals); light red and light blue indicate High-Low and Low-High spatial outliers, respectively. Gray counties show no statistically significant local clustering. Significant clusters are concentrated in a small number of regional pockets, most notably Michigan and Wisconsin.

#### Multi-Season Robustness of Built-Environment Feature Contributions

To evaluate whether the improvement in onset-timing prediction from incorporating built-environment and behavioral residual transmissibility modulation (Fig. 4) reflects a generalizable signal rather than overfitting to a single season, we repeated the residual regression and model comparison across four pre-pandemic influenza seasons (2016–2017 through 2019–2020).

For each season, we compared onset-week prediction error (model minus observed) and rank concordance (weighted Spearman correlation) between the model incorporating connectivity, contact heterogeneity, and school calendar effects, and the expanded model additionally incorporating built-environment/behavioral residual transmissibility (Figure S9).

Incorporating built-environment features improved onset predictions, both in terms of reduced prediction error and increased weighted Spearman correlation, in two of the four seasons evaluated (2016–2017 and 2018–2019). In the remaining two seasons (2017–2018 and 2019–2020), adding built-environment features did not improve, and in the case of 2017–2018 modestly reduced, rank concordance relative to the model without these features.

We interpret this season-dependent pattern as most consistent with behavioral parameters already explaining much of the relevant county-level heterogeneity in onset timing in certain seasons, rather than as evidence against the biological relevance of built-environment predictors. Notably, the 2017–2018 season was atypically severe and prolonged, associated with substantial vaccine-strain mismatch, suggesting that immunological and antigenic factors not included in our current framework may interact with environmental predictors of transmission in season-specific ways. Disentangling these interactions and validating the predictive contribution of built-environment features using explicit train/test splits across seasons or held-out geographic regions, remains an important direction for future work.

**Figure S9:**
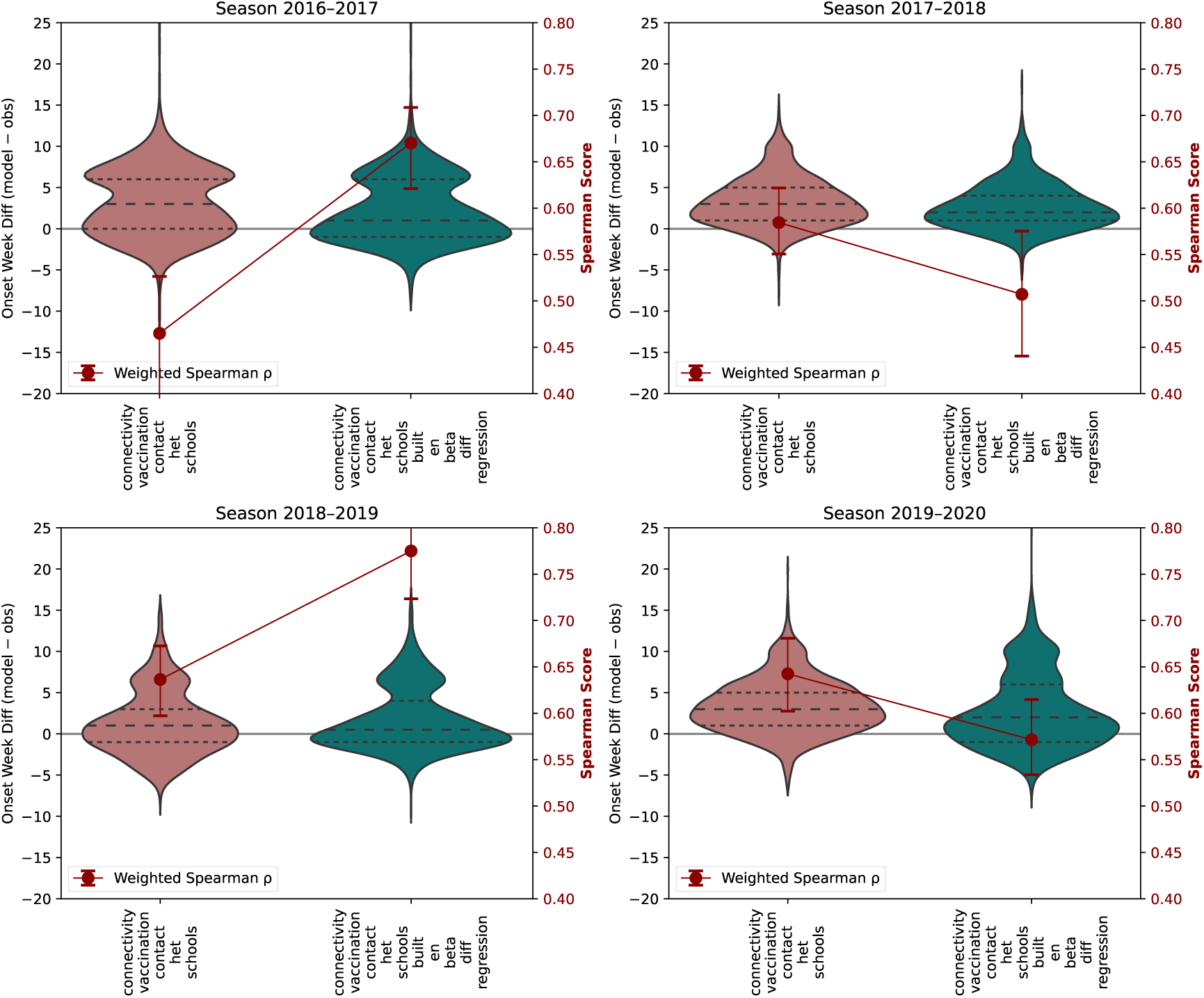
Multi-season comparison of onset-timing prediction accuracy with and without built-environment/behavioral residual transmissibility modulation. Violin plots show the distribution of prediction errors (simulated minus observed onset week) across U.S. counties for the baseline model (connectivity, vaccination, contact heterogeneity, schools) versus the expanded model incorporating built-environment and social mixing, for four influenza seasons (2016-2017 through 2019-2020). Overlaid point estimates and error bars show the population-weighted Spearman correlation (right axis) for each model configuration and season.

##### Validation of Claims-Based Influenza Surveillance Using Wastewater Data

Previous studies have demonstrated strong temporal alignment between these claims-based time series and CDC surveillance data (*63, 67*). We also investigated the alignment of medical claims data with wastewater data from the WastewaterSCAN Dashboard (see Fig S10 and (*63*)).

**Figure S10:**
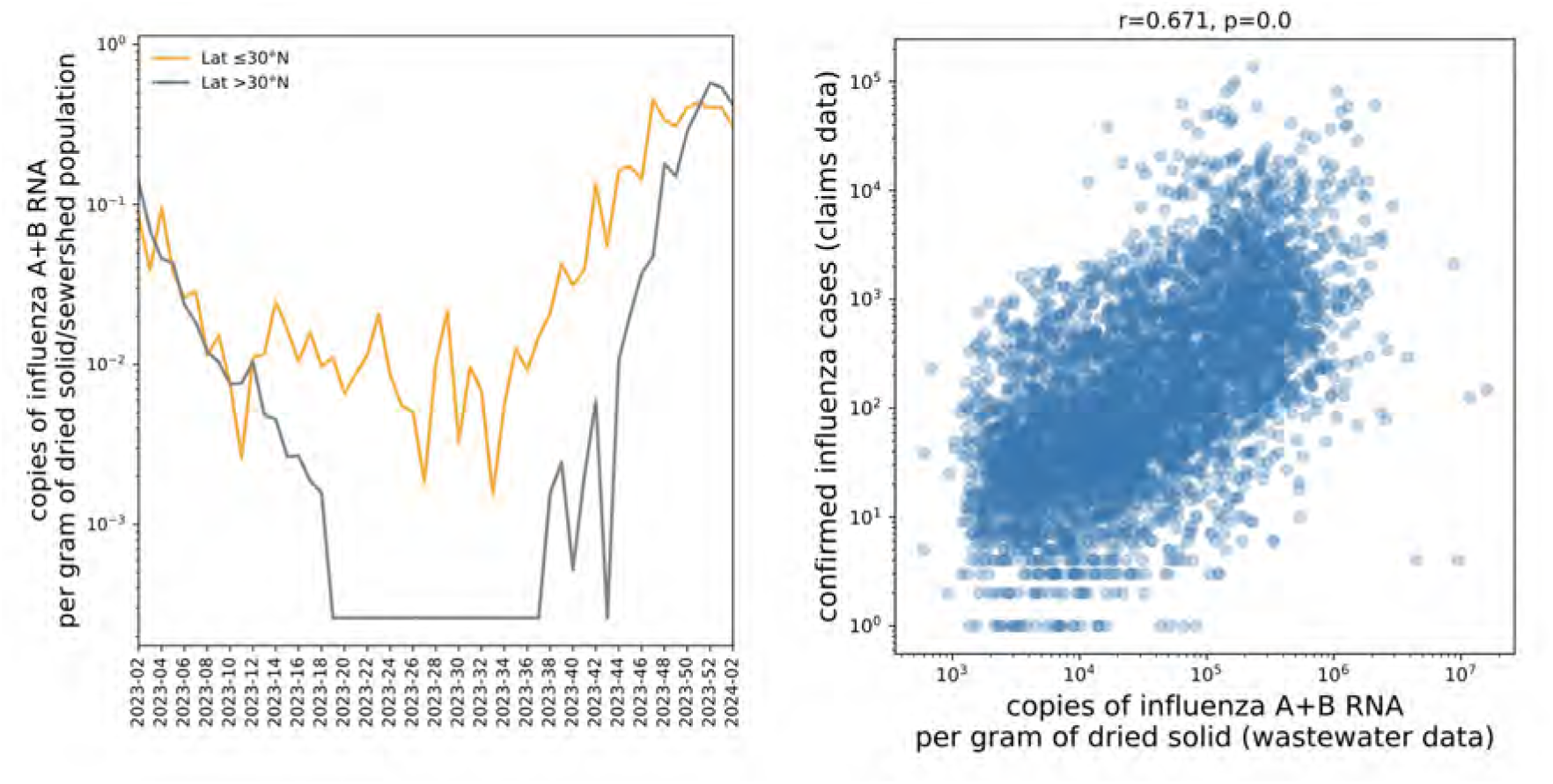
Association between confirmed flu cases from claims data and influenza A and B RNA concentrations in wastewater. (Left) : Time series of copies of influenza A+B RNA per gram of dried solid divided by the sewershed population showing distinct seasonal patterns in northern (Lat > 30^◦^N, gray) versus southern (Lat < 30^◦^N, orange) regions in 2024. (Right) : Scatterplot showing the relationship between weekly confirmed influenza cases from claims data and influenza A+B levels detected in wastewater samples across the 2022–2023 and 2023–2024 respiratory virus seasons. Wastewater data are aggregated across multiple sites at the county level. The observed Spearman correlation (*ρ* = 0.67, *p* < 0.001) suggests a strong positive association between the two datasets.

#### RSV Seasonality analysis

**Figure S11:**
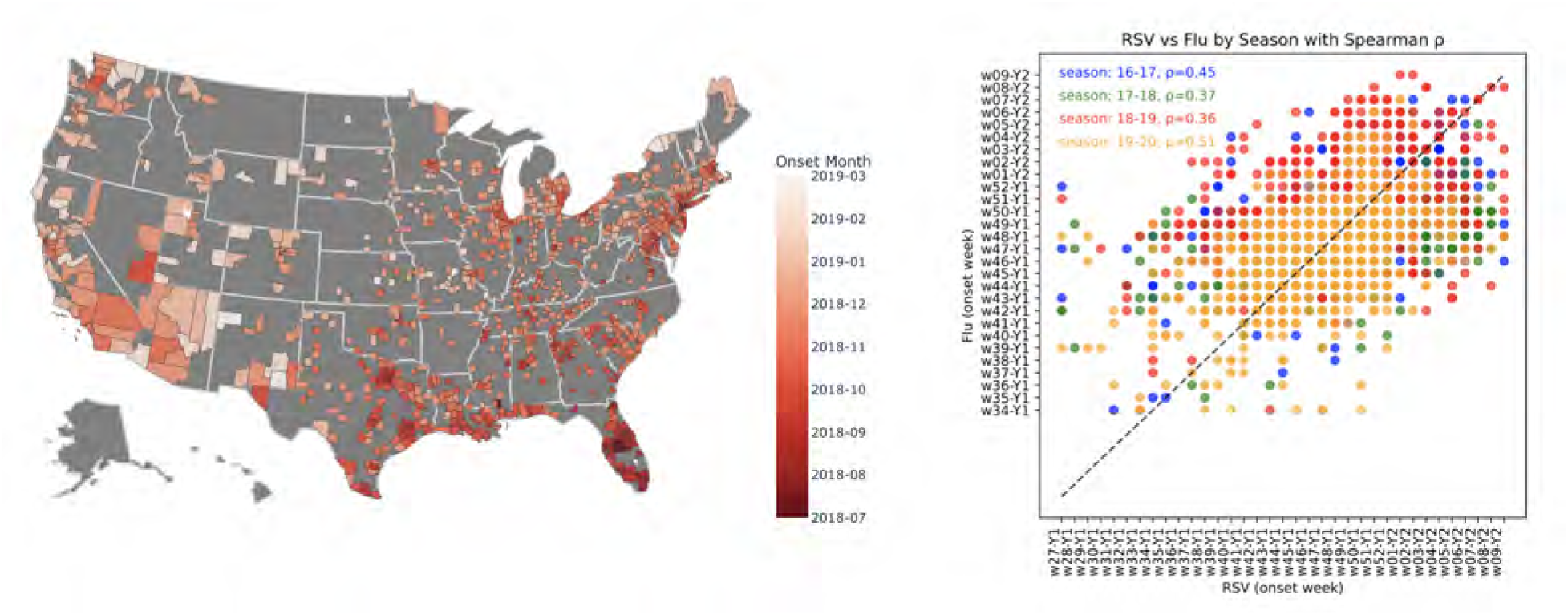
Spatial and temporal relationship between RSV and influenza seasonal onset. (Left): County-level map showing the estimated onset timing of influenza in the 2018–2019 season. (Right): Scatterplot comparing RSV and influenza onset weeks across U.S. counties for four seasons (2106–2020), showing a similar gradient to the influenza geographical spread. Each point represents one county-season, colored by season. Axes reflect epidemiological weeks, with week labels standardized across seasons as wXX-Y1 and wXX-Y2 for weeks occurring in the first or second year of the season, respectively. The dashed diagonal line represents equal onset timing for RSV and influenza. Spearman correlation coefficients (*ρ*) are shown for each season, indicating moderate but variable synchrony in onset timing across respiratory viruses and years.

**Figure S12:**
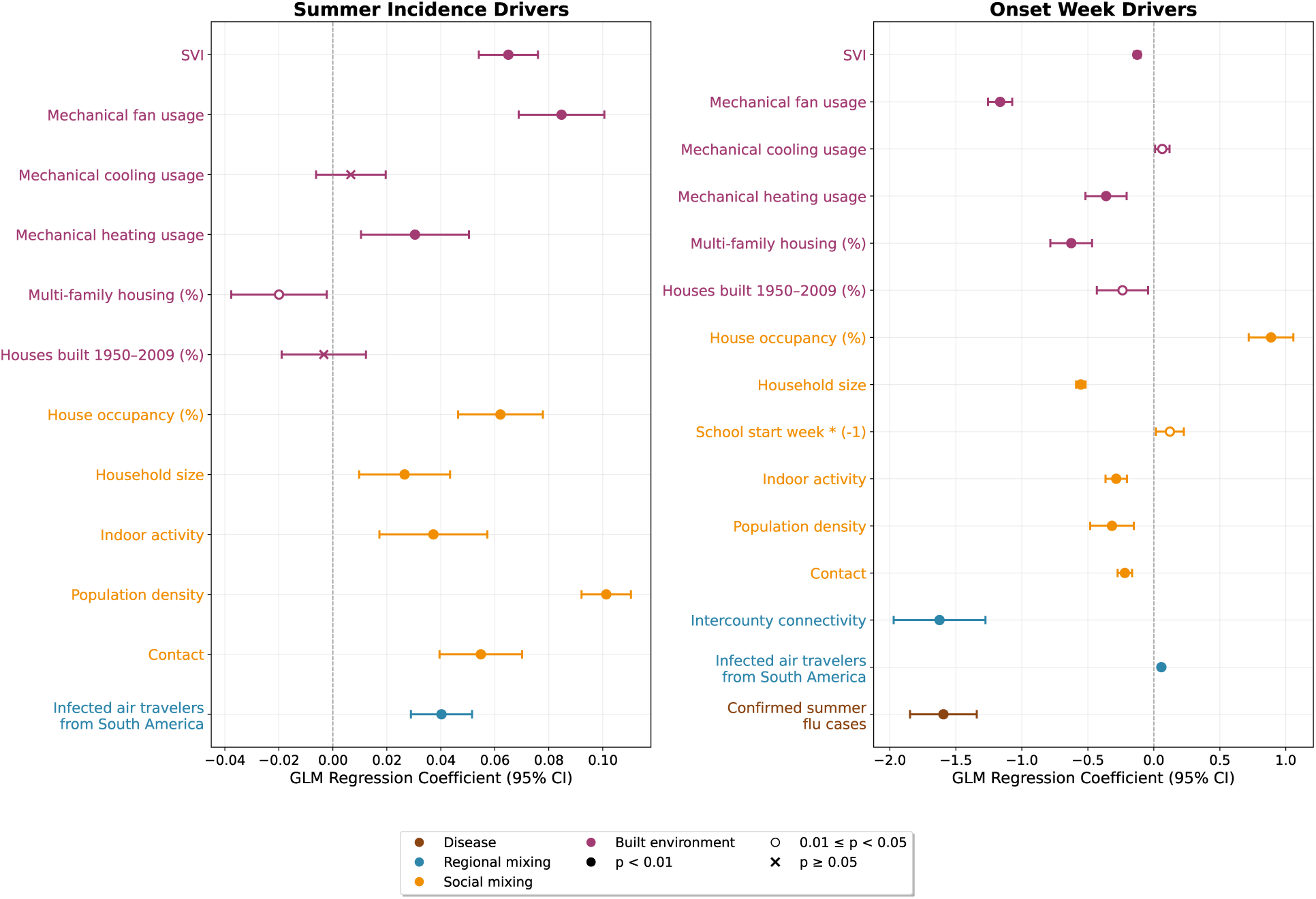
Statistical analysis of factors associated with RSV seasonal dynamics. (a) Standardized regression coefficients and 95% confidence intervals for predictors of summer influenza incidence across U.S. counties, with predictors averaged over weeks 22–26. (b) Standardized regression coefficients and 95% confidence intervals for predictors of epidemic onset timing, with predictors averaged over weeks 29–34. All predictor variables were z-normalized, and models were estimated using generalized linear regression.

#### Environmental Regression Models

To evaluate whether ambient meteorological conditions alone can account for seasonal influenza dynamics, we estimated baseline pooled panel regression models for both summer incidence intensity and epidemic onset timing using purely environmental predictors: feels like temperature, outdoor absolute humidity, total precipitation, and windspeed, with fixed effects for each season.

To formally assess performance, model fits were evaluated using the Akaike Information Criterion (AIC) against our primary behavioral and built-environment models using complete-case data. For summer incidence intensity, the environmental model yielded an AIC of 567, 471.30 compared to 550, 407.00 for the behavioral model, demonstrating a massive reduction in information loss provided by the behavioral framework (ΔAIC = 17, 064.38).

**Table S3:** Environmental Model for Summer Influenza Incidence.

| Variable | Estimate ($\beta$) | Std. Error | *t*-value | *p*-value |
| --- | --- | --- | --- | --- |
| (Intercept) | −5.604558 | 0.002420 | −2316.09 | < 0.001 |
| Feels Like Temperature | +0.071952 | 0.001845 | 39.01 | < 0.001 |
| Outdoor Absolute Humidity | +0.025760 | 0.001948 | 13.22 | < 0.001 |
| Precipitation | +0.009793 | 0.001281 | 7.65 | < 0.001 |
| Windspeed | +0.036924 | 0.001440 | 25.63 | < 0.001 |
| Year Fixed Effect: 2018 | −0.053986 | 0.003250 | −16.61 | < 0.001 |
| Year Fixed Effect: 2019 | +0.077261 | 0.003168 | 24.39 | < 0.001 |

Epidemic onset timing was operationalized as the onset week index (onset idx). The baseline environmental model was estimated on an unbalanced panel of 1,725 counties over 3,871 county-year observations across 4 harvest seasons (2106–2019).

As with summer incidence, the baseline environmental model for epidemic onset (AIC = 17, 030.56) was substantially outperformed by the behavioral and built-environment framework (AIC = 14, 265.36), representing a decisive improvement in explanatory power and information fit (ΔAIC = 2, 765.20).

This confirms that human behavioral patterns and built-environment structures—rather than ambient meteorological conditions alone are the primary determinants of summer influenza dynamics and timing.

**Table S4:**
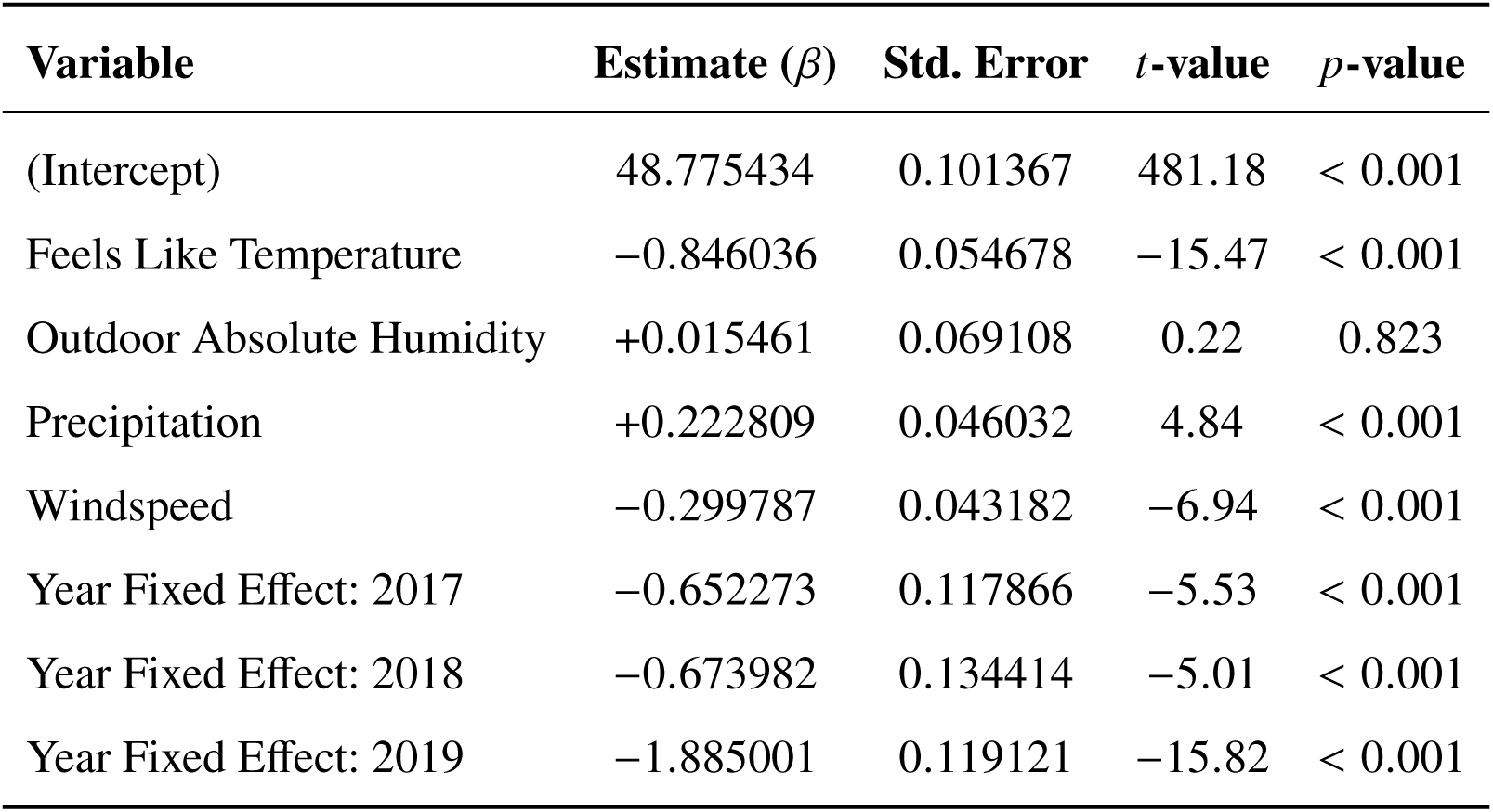
Environmental Model for Epidemic Onset Timing.

#### Epidemiological Model

The age-specific force of infection in county *i* for age group *a* is defined as follows:

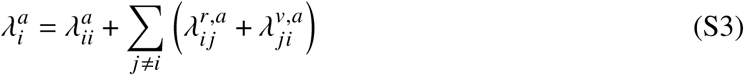

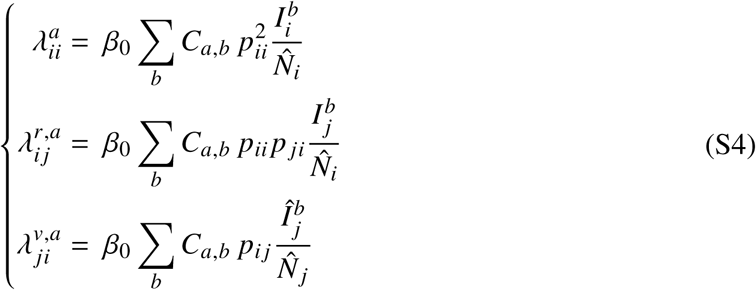

Here *p_i j_* represents the mobility coupling probability between counties *i* and *j* derived from the intercounty connectivity networks, *C_a_*_,*b*_ denotes the time-varying, age-specific contact matrix that switches between school and holiday periods based on county-specific educational calendars. We implemented a two-age-group structure, representing school-age children (5-17 years) and adults (18-65 years) (*8*). The contact matrices are:

During school periods:

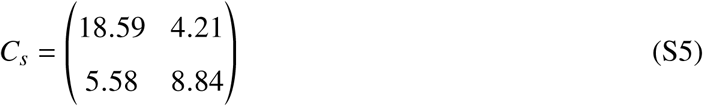

During school holidays:

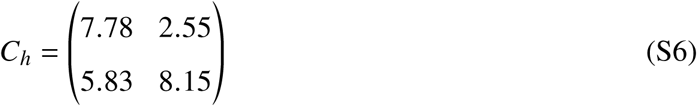

The effective population and infection dynamics account for mobility-driven mixing:

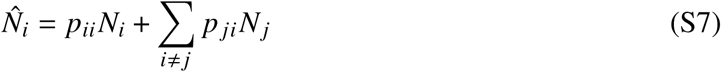

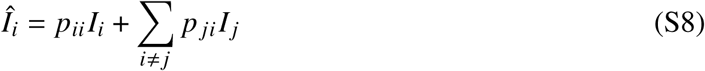

The baseline transmission parameter *β*_0_ was estimated using maximum likelihood estimation applied to national-level influenza incidence data. The calibration period spanned from week 31 (late summer) through week 6 of the following year, covering a complete 27-week epidemic season that captures the full dynamics from onset through peak transmission.

The likelihood function for parameter estimation is defined as:

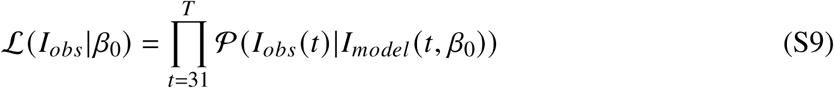

where *I_obs_* (*t*) represents the observed national incidence at week *t*, *I_model_* (*t*, *β*_0_) is the modelpredicted incidence given transmission parameter *β*_0_, and P denotes the Poisson probability mass function.

The maximum likelihood estimate is obtained by:

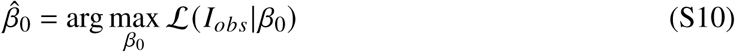

To account for inherent stochasticity in disease transmission dynamics, we performed 50 independent realizations of the model under identical initial conditions.

##### Integration of season and age-specific vaccination rollout

To incorporate season-specific population immunity into the mechanistic model, we constructed weekly, county-level, age-stratified estimates of influenza vaccination coverage by combining two complementary data sources: (i) national CDC weekly cumulative influenza vaccination coverage data by age group (*61*), and (ii) county-level yearly vaccination coverage estimates for the 65+ age group on Medicare enrollees from County Health Rankings (*62*).

We used the 65+ county-level yearly estimates, together with national age-specific coverage ratios, to estimate yearly vaccination coverage for the remaining age groups (5-17 and 18-64) at the county level. Specifically, for each county, we scaled the observed 65+ coverage by the ratio of national coverage in each target age group to national coverage in the 65+ group, yielding a county-level yearly coverage estimate for each age group consistent with observed national age patterns. We then used the national CDC weekly cumulative coverage data to characterize the within-season temporal shape of vaccination uptake for each age group. This profile captures the characteristic rollout of vaccination coverage from the start of the campaign (July) through its plateau later in the season.

#### Linking Regression Frameworks to Residual Transmissibility Analysis

To systematically evaluate the predictors of seasonal influenza dynamics, we implemented a twostage analytical framework. In the first stage, candidate covariates spanning susceptibility, built environment, social mixing, regional connectivity, and observed disease dynamics were evaluated using statistical regression models predicting summer incidence and epidemic onset timing. This step identified significant empirical predictors without imposing prior mechanistic constraints.

In the second stage, significant covariates were integrated into the mechanistic SEIR metapopulation model via one of two distinct pathways:

1. Direct Mechanistic Parameters: Variables with direct, well-established structural representations within transmission dynamics, specifically population density, school reopening timing, intercounty human mobility, contact heterogeneity, and confirmed summer influenza cases, were incorporated directly as mechanistic inputs into the SEIR differential equations.
2. Residual Transmissibility Predictors: Variables that demonstrated statistical significance but lacked a direct, closed-form structural equation within the standard SEIR formulation (such as mechanical ventilation, multi-family housing, and indoor activity) were evaluated in a second-stage residual transmissibility regression. These variables modulate county-level transmission rates (*β*) indirectly, capturing unmeasured micro-environmental and behavioral heterogeneity.

Table S5 provides a comprehensive summary of statistical significance across primary regression models and their explicit structural or indirect roles within the mechanistic SEIR metapopulation framework.

**Table S5:**
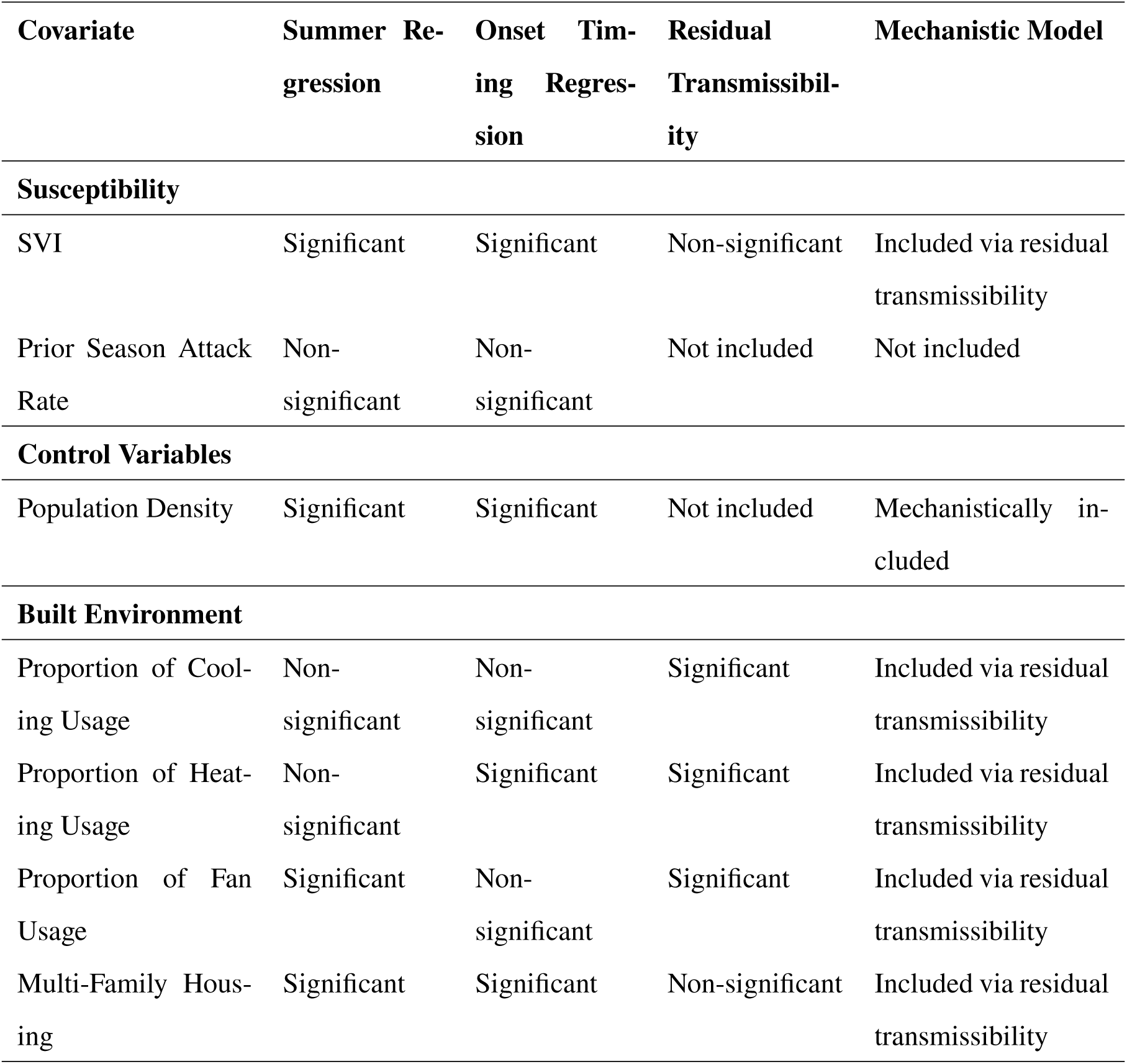
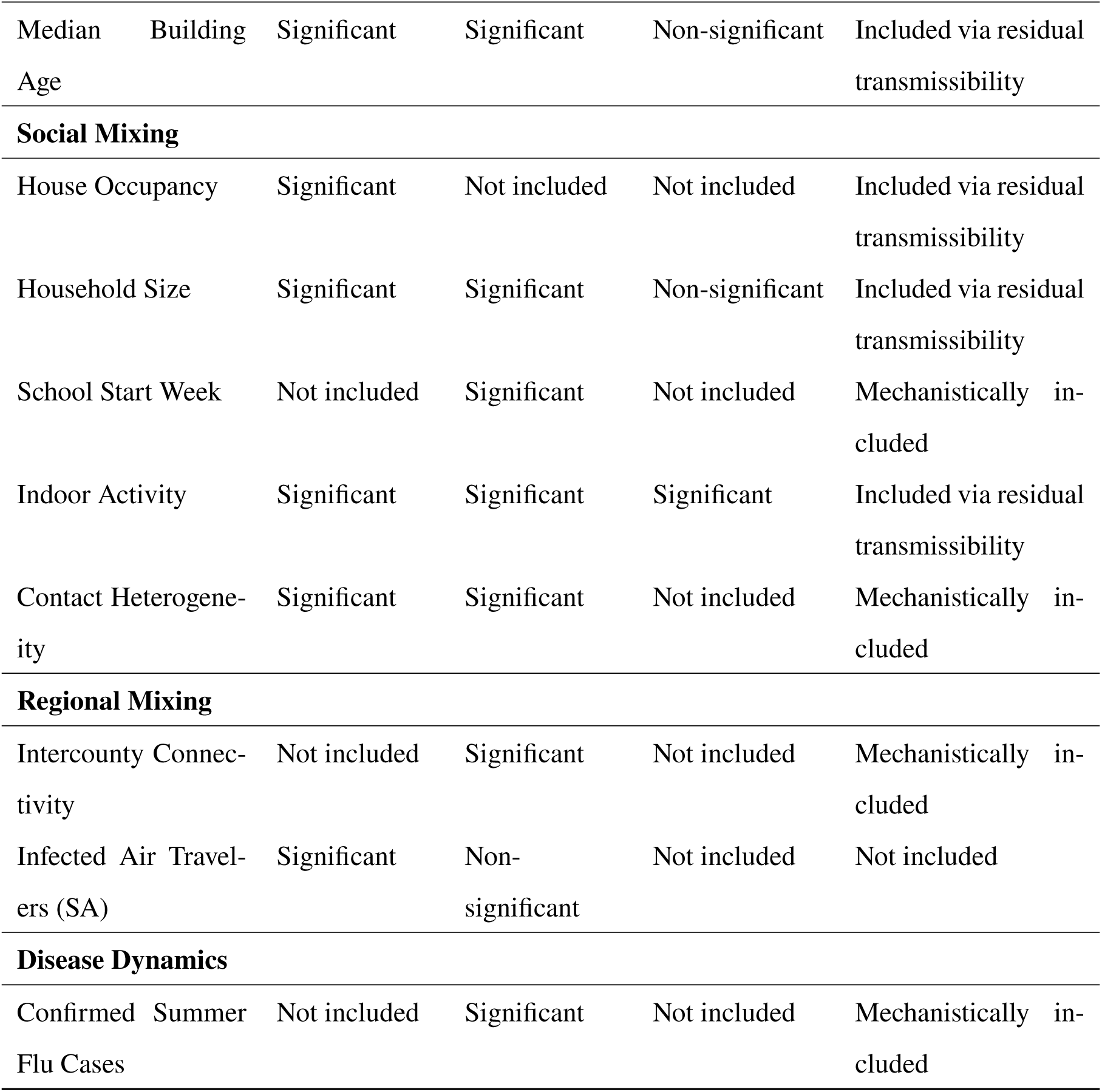

#### Estimated Vaccination Rollout in the US

##### Vaccination Policy Scenarios

**Figure S13:**
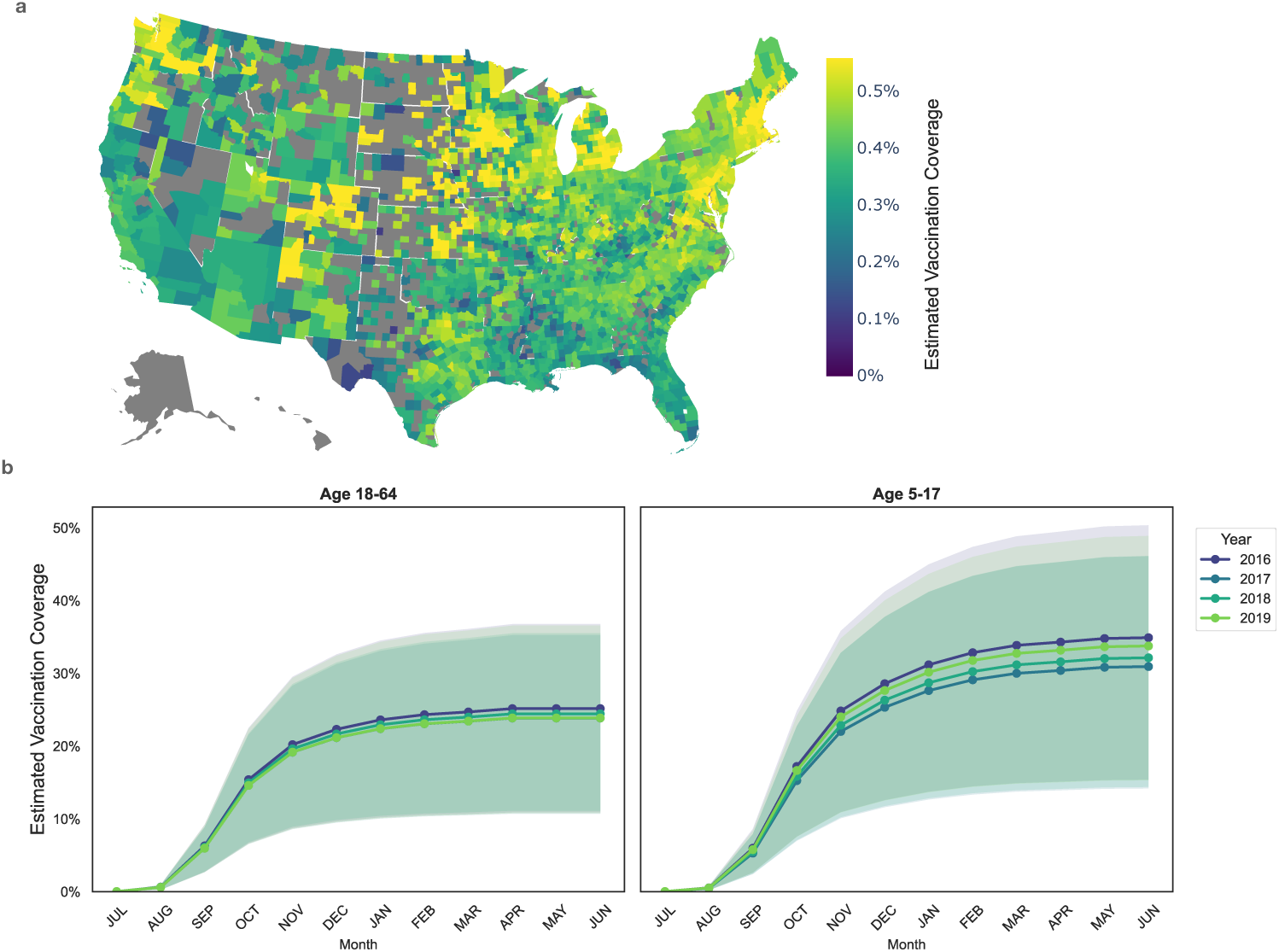
Estimated Vaccination Rollout in the US. (a) Map of estimated vaccination coverage by US county in 2018. (b) Monthly estimated vaccination coverage by year and age group. Median and 95% CI across counties.

**Figure S14:**
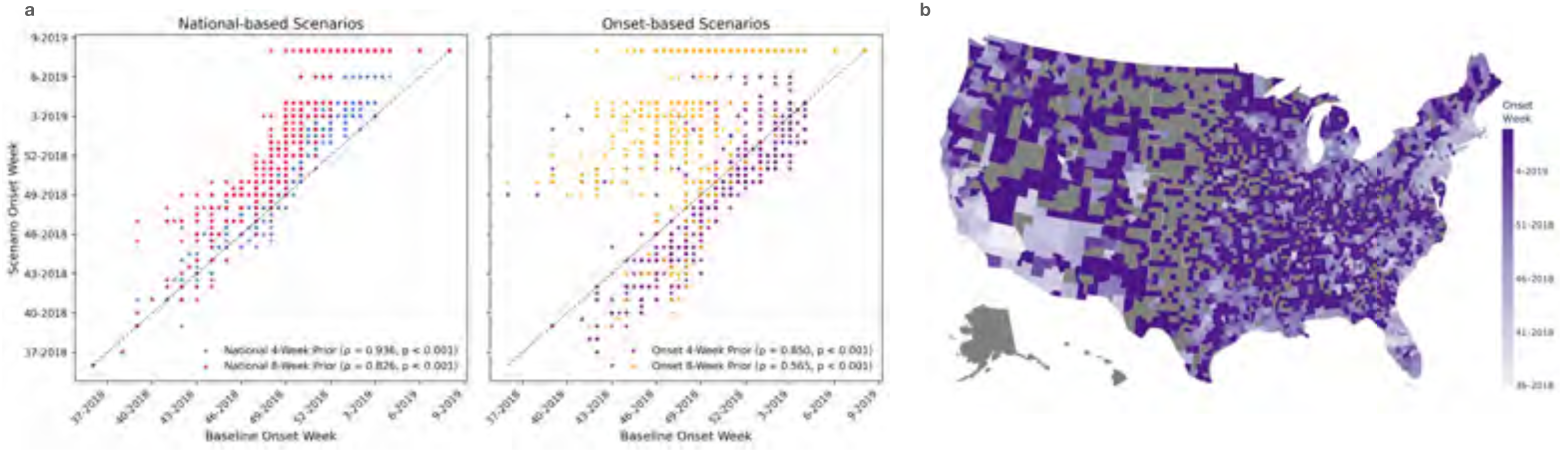
Impact of timing and geographical targeting on vaccination intervention effectiveness. (a) Scatter plots comparing the baseline epidemic onset week against the scenario onset week for National-based (left) and Onset-based (right) interventions. Spearman correlation coefficients (*ρ*) and p-values denote the geographical shift in onset timing. (b) Map of the continental United States displaying the modeled epidemic onset week by county for the Onset 4-Week Prior scenario.

**Figure S15:**
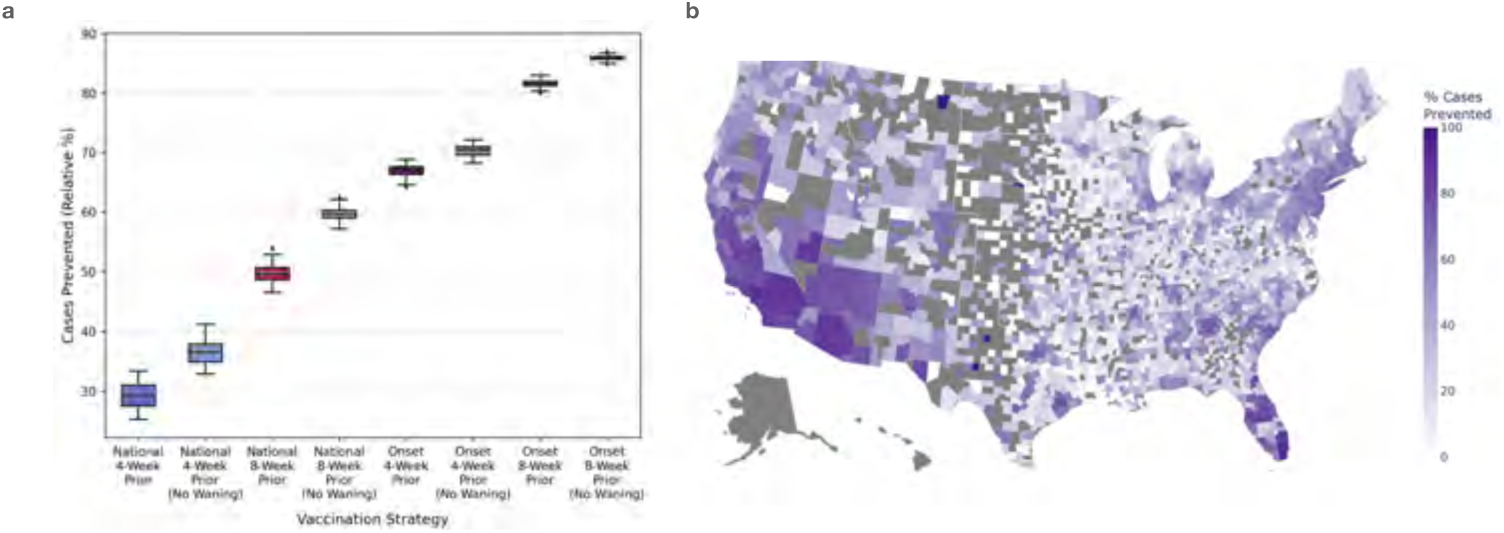
Sensitivity analysis vaccination intervention effectiveness. a) Boxplots displaying the relative percentage of cases prevented across four vaccination strategies: National 4-Week Prior, National 8-Week Prior, Onset 4-Week Prior, and Onset 8-Week Prior. Results are displayed for scenarios modeled both with and without waning immunity. (b) Map of the continental United States illustrating the relative percentage of cases prevented at the county level for the Onset 4-Week Prior scenario

## Notes

### Competing Interest Statement

The authors have declared no competing interest.

### Author Declarations

This study was reviewed by the Georgetown University Institutional Review Board and determined to be exempt from full IRB review under [Exemption Category, e.g., 45 CFR 46.104(d)(2)] (Protocol # STUDY00005121) or has been approved under expedited review (Protocol # STUDY00002324, STUDY00003041).

